# Impact assessment of vaccine-related negative news and incentive measures on vaccine hesitancy in Hong Kong

**DOI:** 10.1101/2025.02.21.25322646

**Authors:** Yifan Chen, Yang Ye, Hsiang-Yu Yuan, Qingpeng Zhang

## Abstract

Vaccine hesitancy underscores the critical need to quantify how diverse factors shape vaccine uptake. We develop a social-epidemiological transmission model with a game-theoretic imitation mechanism and payoff-driven risk perception to assess the impacts of vaccine-related negative news and incentive measures on COVID-19 vaccine uptake during the pandemic. By fitting our model with real-world data from Hong Kong, we reveal that the negative news drastically impeded vaccination efforts. Scenario analyses suggest that, without incentive measures, the projected fifth wave of COVID-19 in Hong Kong (from December 31, 2021, to October 15, 2022) would have infected 97.3% of the population, resulting in an estimated 48,892 deaths. Both model simulation and real-world data demonstrate that incentive measures have successfully encouraged vaccine uptake and saved approximately 39,073 lives. However, we found that vaccine willingness declined rapidly after the incentive measures discontinued, implying limited benefits in mitigating the effect of negative news in the long run. This study also highlights the need for booster doses in the face of the immune escape of the Omicron variants. Our model offers data-driven insights into the interplay between negative news, vaccine hesitancy, and incentive measures, shedding light on the effective preparation for emerging infectious disease outbreaks.

## Introduction

Vaccine hesitancy has been among the top ten global health threats identified by the World Health Organization (WHO)^1^. The ongoing mutations of COVID-19 highlight the need to address vaccine hesitancy, as it can lead to repeated infections and elevate the risk of developing Long COVID^2,3^. Vaccine hesitancy extends beyond COVID-19. Recent WHO statistics indicate that vaccination coverage rates for measles, diphtheria-tetanus-pertussis (DTP), and human papillomavirus (HPV) fall short of established targets^4^. This reluctance to vaccinate undermines the United Nations’ Sustainable Development Goal (SDG) of healthy lives and well-being for all at all ages^5^. Addressing vaccine hesitancy is essential for achieving better health outcomes globally.

Vaccine hesitancy is multifactorial, spanning psychological, social, and structural dimensions^6^, which can be conceptualized through the 5C scale^7^: *confidence*^8–12^ (trust in vaccine safety and efficacy, delivery systems, and governance), *complacency*^13–15^ (perceived low risk of disease or lack of urgency to vaccinate), *constraints*^16^ (barriers such as cost, access, or logistical limitations), *calculation*^17–19^ (active information seeking), and *collective responsibility*^20,21^ (the willingness to protect others by one’s own vaccination by means of herd immunity).

Notably, negative news directly erodes confidence (e.g., safety scandals^10,11,22^) and fuels calculation (e.g., misinformation-driven risk overanalysis^23^). Historical misuse of medical and vaccine research during the colonial era in Africa continues to affect COVID-19 vaccination rates on the continent^24^. Negative news has stronger effects on public trust than positive news, often resulting in greater public engagement^25–27^ and the proliferation of conspiracy theories and misinformation^28^. Negative news thrives in polarized environments: right-wing or conservatives exhibit relatively higher susceptibility to conspiracy theories and misinformation^18,29–35^, often exacerbating vaccine hesitancy under the governance of right-wing parties or the claim of prominent anti-vaccine activists. Conversely, incentive measures primarily address constraints but may indirectly affect confidence through institutional trust signaling. In Europe, cash incentives were effective in boosting vaccination rates^36^. Interestingly, smaller cash rewards positively affected vaccination rates more than higher cash rewards^37^. However, several studies have shown that negative news regarding the Oxford AstraZeneca vaccine did not affect its uptake in the UK and Denmark^38–40^, and economic incentives did not increase overall vaccination rates at all in the United States^41,42^. Additionally, a previous study has indicated that in Hong Kong, the incentive measures have little impact on encouraging older adults to get vaccinated^43^.

The COVID-19 epidemic in Hong Kong provides an ideal case study, offering a real-life experimental setting to assess the impact of vaccine-related events on vaccine hesitancy. During the pandemic, the public willingness to get vaccinated declined due to vaccine-related negative news, including packaging defects, serious adverse events after vaccination, and reduced information about deaths possibly linked to vaccination^44–46^. Subsequently, various incentive measures were introduced, including material gifts, economic rewards, and vaccination leave^47^. While many surveys have examined the effects of various factors on vaccine hesitancy in Hong Kong^48–52^, predefined surveys often fail to capture all relevant factors and events, and limited sample sizes often lead to selection bias. Thus, understanding these vaccine-related events is crucial for addressing vaccine hesitancy effectively.

In this study, we aim to leverage real epidemiological data to assess the impact of vaccine-related negative news and incentive measures on vaccine hesitancy in Hong Kong. We develop a social-epidemiological model to account for key events affecting vaccine uptake. With this model, we conduct scenario analyses to evaluate how these elements affect the number of infections and deaths.

## Results

To evaluate the impact of vaccine-related events on vaccine uptake in Hong Kong, we used a deterministic, SEIRD-like model-based framework to model COVID-19 epidemic dynamics and daily vaccination administration (see Supplementary Fig. 1). This model incorporated time-varying daily vaccination rates modulated by risk perception dynamic, where vaccine uptake is quantified through a composite payoff function comprising three synergistic components: infection risk and mortality risk reduction after vaccination, and vaccine-related events including negative news and incentive measures.

### Model-based infection-vaccination dynamics and event-driven behavioral shifts

We explored the impact of vaccine-related negative news and incentive measures on infection-vaccination dynamics in Hong Kong. To do so, we first fitted the social-epidemiological model to daily reported infected cases and count of vaccination in Hong Kong. The study period spanned from January 24, 2020, to October 15, 2022, which was delineated into three phases based on vaccine availability (see the Methods for more details). Our model successfully reproduced the epidemic dynamics and daily vaccination trends across the study period, with reasonable consistency between observed data and model fits (see Fig. 1 for fits; Supplementary Figs. 2-12 for full model diagnostics). Of note, fits were poorer for the booster vaccination phase due to the limitations of segmented modeling (Fig. 1c). These segments were driven by two key events: E1 (February 26, 2022): a sudden surge in reported infected cases because the government included positive RAT (rapid antigen tests) cases in official statistics; E2 (May 23, 2022): the emergence of a new epidemic wave. Nonetheless, the consistency of fits across multiply-independently sampled chains allowed us to reliably assess the impact of vaccine-related events on vaccine uptake in Hong Kong.

**Figure 1.**
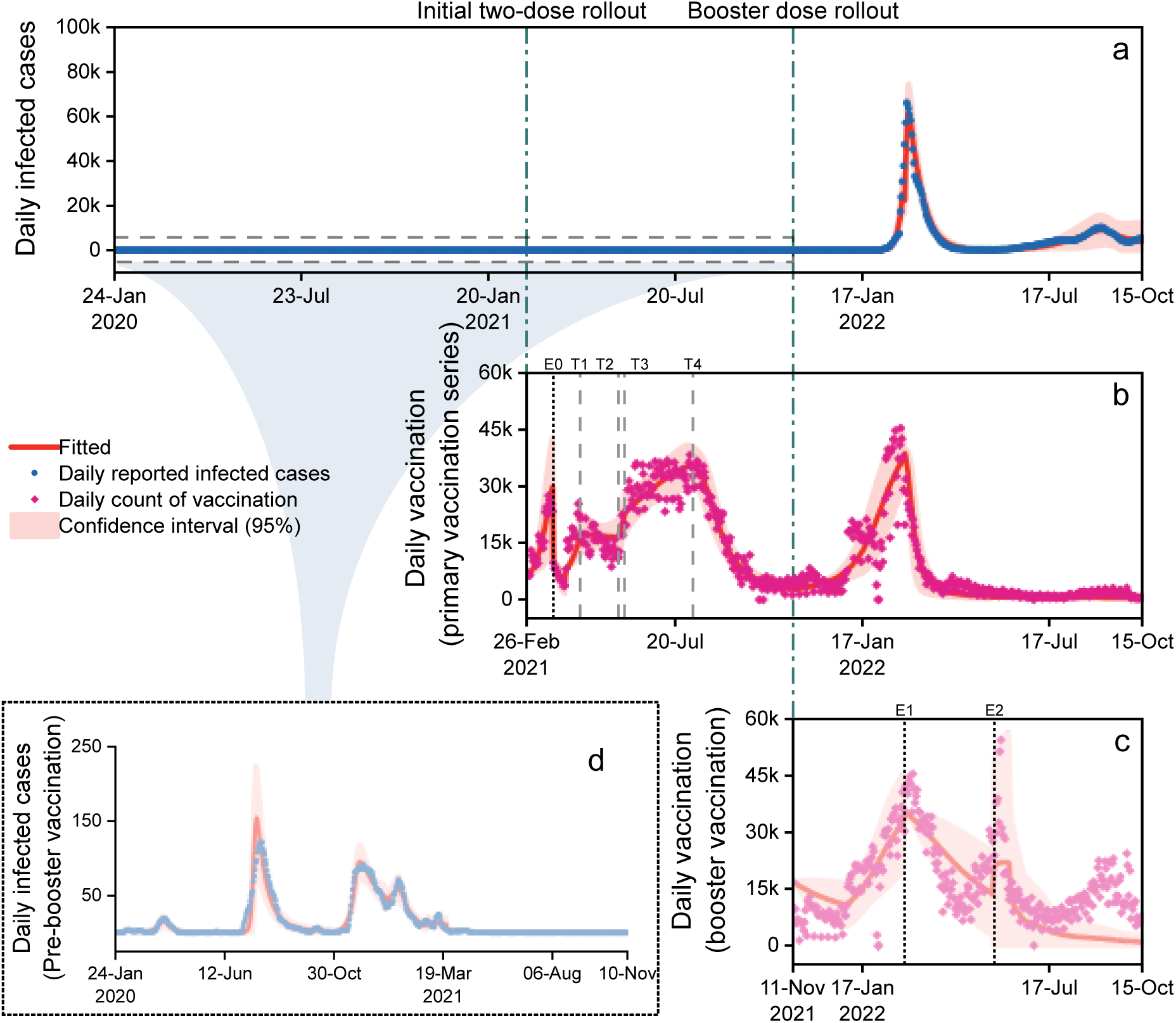
COVID-19 epidemic dynamics and vaccination in Hong Kong (Jan. 2020-Oct. 2022). (a) Daily new reported infected cases (blue circles) and model-fitted transmission trend (red line; 95% CI shaded); (b)-(c) Daily count of vaccination (magenta diamonds) and model-fitted vaccination rates; (d) Zoomed-in view of the marked period in panel a (dashed shaded grey box) showing detailed fitting. Black dotted line in panel b and c show timepoints of fitting segmentation (E0-E2). Gray dashed lines in panel b show timepoints of four critical events (T1-T4).

We used the estimated payoff gains to quantify how screened critical events influenced people’s willingness to get vaccinated (Table 1). The analysis revealed that negative news substantially reduced the perceived benefits of vaccination. Notably, the vaccine packaging defects (E0) led to a reduction in the payoff gain, decreasing it by 232.4% (95% CI: 158.3%-306.5%). This event had a markedly greater impact on vaccination hesitancy compared to the other two events, the serious adverse events after vaccination (T1: 100.7%, 95% CI: 76.6%-124.9%) and reduced information about deaths possibly linked to vaccination (T3: 77.1%, 95% CI: 40.0%-114.2%).

**Table 1.** The impact of different events on vaccine uptake.

| Timepoints of events | Events | Potential 5C scale | Payoff gains for primary vaccination series |
| --- | --- | --- | --- |
| 2021.03.24 | E0: Vaccine packaging defects | Confidence | -130.8(-232.4%) |
| 2021.04.19 | T1: Serious adverse events after vaccination | Confidence | -71.9(-100.7%) |
| 2021.05.26 | T2: Government and business incentive measures | Constraints, confidence | 64.6(90.6%) |
| 2021.06.01 | T3: Reduced information about deaths possibly linked to vaccination | Confidence, calculation | -55.0(-77.1%) |
| 2021.08.08 | T4: End of the incentive measures | Constraints | -67.5(-94.6%) |

In response to a slow vaccine rollout of COVID-19, the Hong Kong government and local private sectors introduced several incentive measures (T2) to encourage vaccination, including lucky draws and cash bounties, which increased the benefits of receiving primary vaccine doses. However, their effect was temporary, as evidenced by a reduction in payoff gain once these reward programs concluded (T4: 94.6%, 95% CI: 66.1%-123.1%).

### Impact of negative news mitigation on vaccination uptake and epidemic trajectories

Our simulations demonstrated how mitigating the effects of negative news can alter vaccination trends and ultimately reduce the public health burden of COVID-19. We simulated the transmission dynamic and daily vaccination rates under four scenarios without incentive measures (Fig. 2): (1) negative news only (the scenario with negative news but without incentive measures; the yellow curve); (2) no news & incentives (the scenario with neither negative news nor incentive measures; the teal curve); (3)-(4) mitigating 50% and 25% of the effects of negative news without incentive measures, respectively (purple and dark brown curves). These simulations were compared to the actual scenario (red curve) to assess how different levels of negative news influence vaccination rates and transmission dynamics.

**Figure 2.**
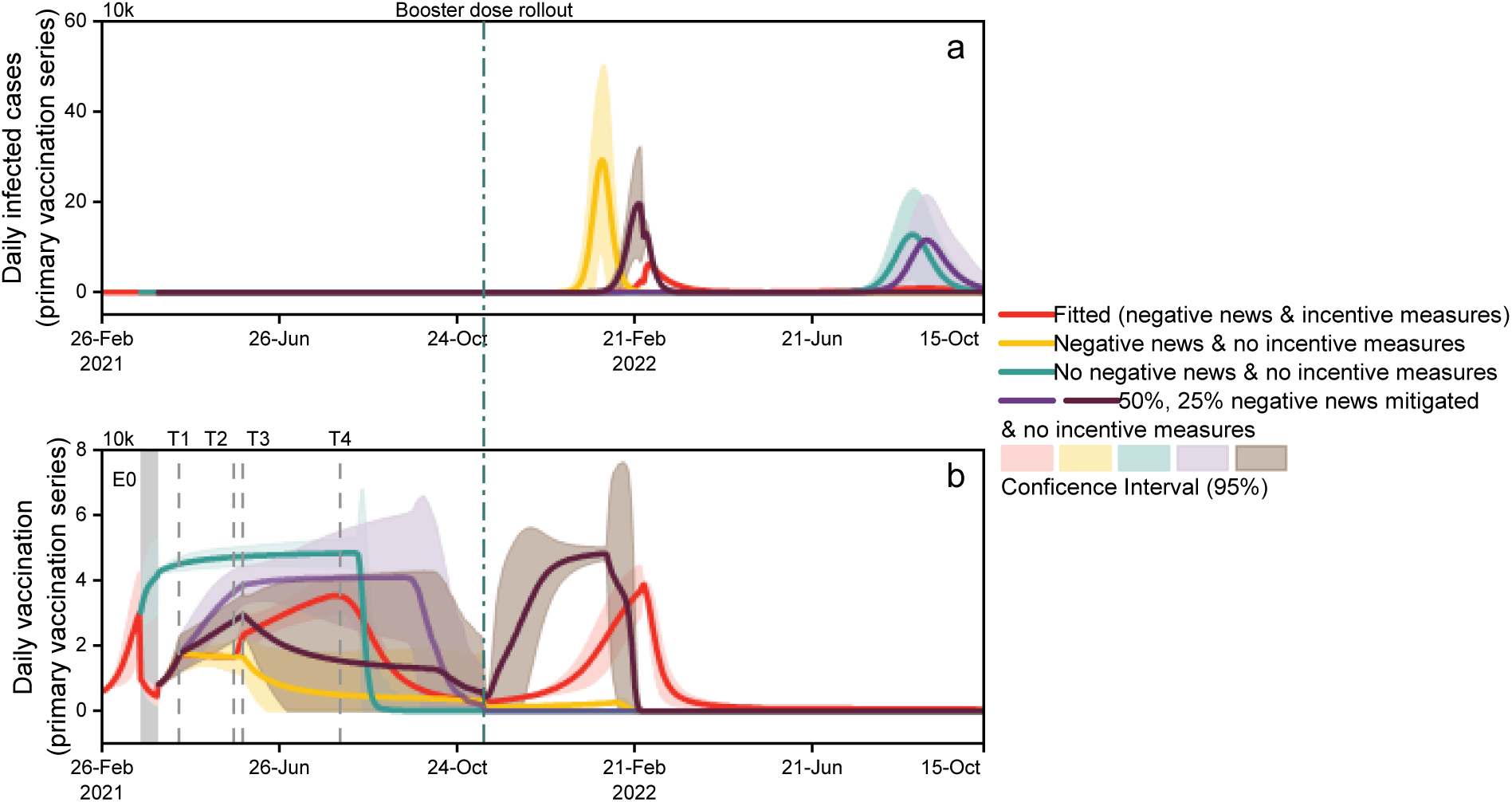
COVID-19 epidemic dynamics and vaccination over time under scenarios without incentive measures (primary vaccination series phase). (a) The transmission trend; (b) Vaccination rates. The solid curves and bands represent the mean of fitted or simulated results, and 95% confidence intervals, respectively. The simulated results of scenarios are represented in yellow, teal, purple and dark brown, respectively.

Comparing the “negative news only” scenario with the actual scenario, we found that, without any incentive measures, the daily vaccine uptake could be much lower than that of the actual scenario. Upon the arrival of the Omicron variant, the epidemic would peak earlier than anticipated, with peak daily infections reaching 3.81 times (95% CI: 1.11-6.50) the actual peak. Without the negative news, daily vaccinations would continue to rise until reaching maximum capacity and remain steady until mid-August, achieving the first primary doses vaccination rate of 99.7% by booster dose rollout—56.5% higher than actual figures.

With universal primary vaccination doses, the Omicron wave in mid-January 2022 would be averted, substantially reducing infection and mortality rates. However, by late July 2022, a more serious wave of outbreaks would emerge caused by the Omicron mutant strain sub-lineages BA.4/BA.5^53^. In this scenario, the projected cumulative number of infections was approximately 4.02 million, which was 2.19 million more than the fitted actual figure. Despite the increase in infected cases, an estimated 5,284 lives would be saved by the study’s conclusion.

Additionally, we found that without incentive measures, the government must mitigate at least 50% of the negative news impact to achieve high coverage (99.7%) of the population receiving primary doses by booster dose rollout. As a result, the fifth wave of outbreak in Hong Kong (in early 2022) could have been prevented.

### The synergistic effect of introducing incentive measures and mitigating negative news

Our simulations demonstrated that combining partial mitigation of negative news with incentive measures substantially enhanced vaccination coverage, but booster dose incentives were critical to countering Omicron immune evasion. We examined how the proper mitigation of negative news may affect the daily infection and vaccination rates under three scenarios with incentive measures (Fig. 3): (1) incentive measures only (the scenario without negative news but with incentive measures; the yellow curve); (2)-(3) mitigating 10% and 5% of the impact of negative news with incentive measures, respectively (since incentive measures can compensate for approximately 50.9% of the impact of negative news, shown in Table 1).

**Figure 3.**
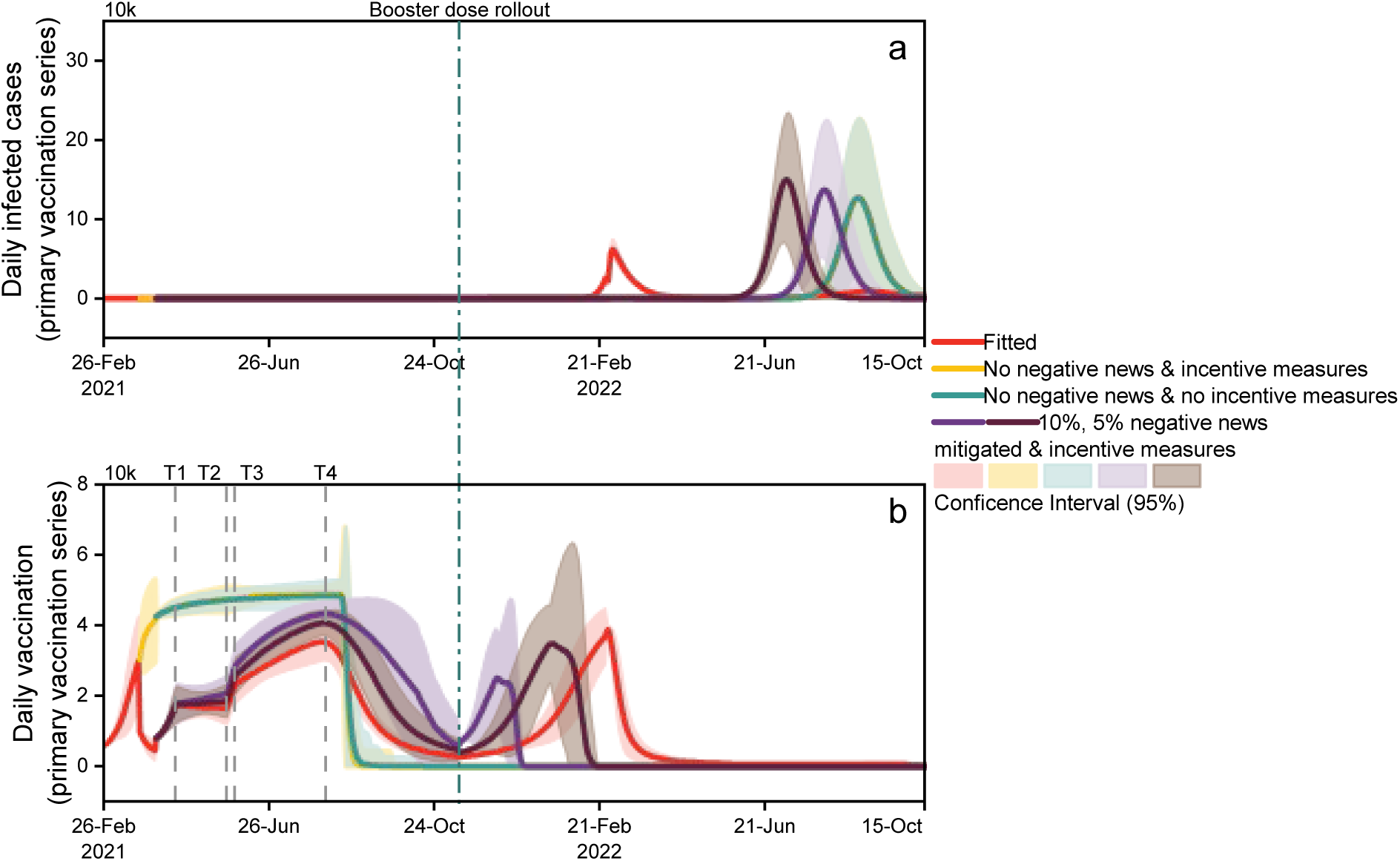
COVID-19 epidemic dynamics and vaccination over time (primary vaccination series phase). (a)-(b) The transmission trend and vaccination rates under four scenarios. The solid curves and bands represent the mean of fitted or simulated results, and 95% confidence intervals, respectively. The simulated results of scenarios are represented in yellow, teal, purple and dark brown, respectively.

In both the scenarios “no news & incentives” and “incentive measures only”, the daily vaccine uptake shared a similar pattern regardless of the incentive measures (as shown by the yellow and teal curves in Fig. 3). Importantly, introducing incentive measures could effectively reduce vaccine hesitancy so that as little as 5% negative news mitigation was sufficient to prevent the fifth outbreak. However, merely increasing the vaccination rate of the two primary doses was insufficient to prevent the future outbreak due to the immune escape of the Omicron variants and the low booster dose vaccine uptake (Fig. 3a, Aug. 2022).

To address this challenge, we simulated how the incentive measures for booster doses to contain the future outbreak. The results showed that if small- to medium-level (25% to 50%) incentive measures were introduced for booster doses after the primary doses vaccination program, the future Omicron-led outbreak could be curbed, potentially reducing infections by 6.4 million and saving 4,748 lives throughout the study period, compared with a scenario without any incentive measures for booster doses (Fig. 4).

**Figure 4.**
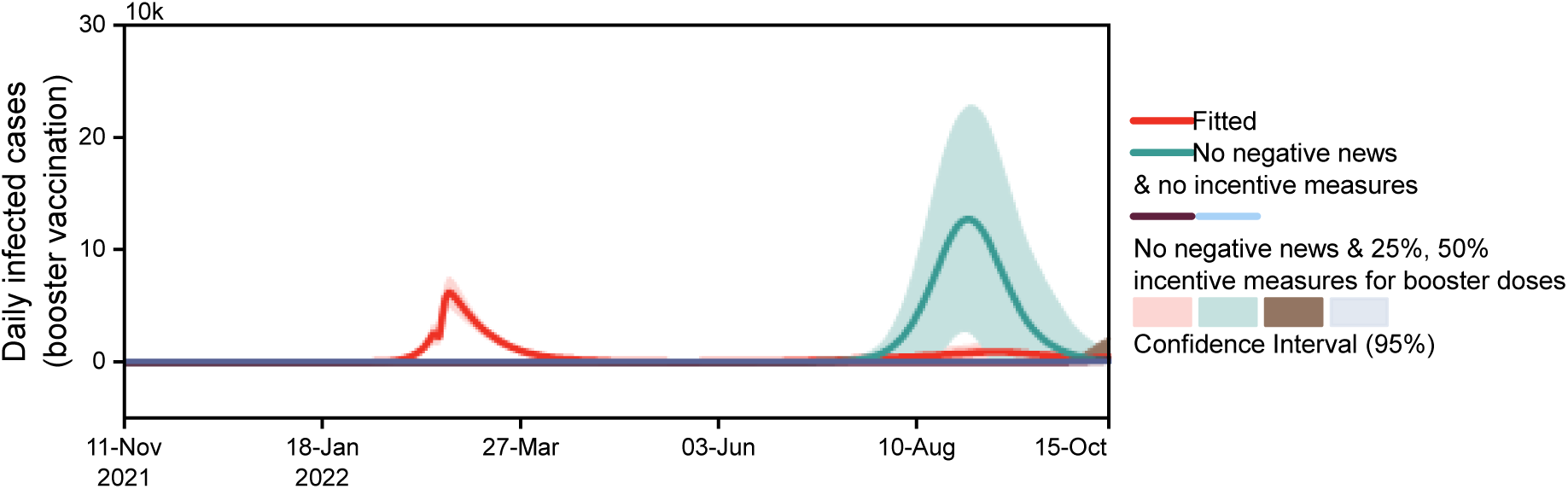
COVID-19 epidemic dynamics over time (booster vaccination phase). The transmission trend under three scenarios. The solid curves and bands represent the mean of fitted or simulated results, and 95% confidence intervals, respectively. The simulated results of scenarios are represented in teal, dark brown and light blue, respectively.

## Discussion

In our study, we fit real epidemic dynamic data to understand how vaccine-related events influenced vaccine uptake in Hong Kong. Our research offers evidence and insights, highlighting the positive synergistic effect of managing negative news and introducing incentive measures to achieve herd immunity. We recommend that only modest incentive measures for booster doses are sufficient to control outbreaks caused by immune escape.

This study provides data-driven evidence for previous survey studies^49–51,54^, assessing the importance of specific vaccine-related events, including the impact of negative news and incentive measures on vaccine uptake. Negative news may reduce vaccination willingness through dual pathways: *confidence* (reports of adverse events and packaging defects reduced trust in vaccine safety, which is a primary concern across all age groups in Hong Kong^49^) and *calculation* (reduced information about deaths prolonged risk-benefit analysis). Negative news may directly undermine trust in the government and contribute to the spread of misinformation and rumors, which has been shown further to amplify vaccine hesitancy through compounded psychological and social mechanisms^55^. Mitigating the effects of negative news by 50% can achieve high coverage of the first two primary doses and save costs on incentive measures, allowing resources to be reallocated toward increasing vaccine supply, expanding healthcare capacity, and supporting economic growth. Notably, previous research has shown that providing information about vaccine risks does not influence willingness to vaccinate^56,57^. Our results suggest that the timely, proper, and transparent publicity of pandemic and vaccine safety-related information could boost vaccination rates by mitigating misinformation originating from negative news. Our findings show that incentive measures may compensate 50.9% of negative news’s impact through *constraints* to boost vaccine uptake. Ultimately, it will aid in the faster achievement of herd immunity and minimize unnecessary infections and deaths. However, their effects are short-term, and vaccine hesitancy remains slightly increased after the incentive measures end.

Our results shed light on the fact that even with high vaccine coverage of first primary doses, neglecting booster doses could lead to a stronger outbreak of Omicron in Hong Kong. The principle underlying the outbreak is the accumulation of susceptible individuals and the relaxation of PHSMs, which aligns with our previous research on respiratory infectious diseases^58,59^. In addition, the Omicron BA.4/BA.5 resurgence despite high primary dosing further highlights *complacency*’s role in the low infection phase, which implicated waning variant vigilance. Thus, when addressing future virus spread and vaccine promotion, the government should not only focus on current epidemic dynamics but also consider virus characteristics to develop seamless and comprehensive strategies. Fortunately, we observed that minimal incentive measures for booster doses can prevent this outbreak. The synergistic effect of combining incentive measures and negative news mitigation indicates a cost-effective approach to promoting vaccination programs during an epidemic.

Our research has limitations. First, our study’s timeframe was limited to data available up to October 15, 2022, due to the availability of Google Mobility data. It is important to identify alternative public human mobility data for future epidemic surveillance and modeling. Second, this study did not explore how negative news and incentive measures affect different socioeconomic groups. A study has indicated that in Hong Kong, incentive measures have little impact on encouraging older adults to get vaccinated^43^. Our model is flexible and can be easily extended to account for the composition of different subpopulations, aiding the formulation of more targeted policy measures. It is important to explore age-specific responses to these key events given representative survey data. Third, our study focused on data from Hong Kong. To expand our research to other regions, we would require relevant epidemic data and event information from each area. However, some regions lack available data, which limits our study to Hong Kong. Nonetheless, our model can be easily adjusted to evaluate vaccine-related events in different regions, helping them develop vaccination strategies suited to their specific needs.

In summary, our research assessed the effects of negative news and incentive measures on vaccine willingness during the COVID-19 crisis. We highlighted the importance of mitigating the impact of negative news and the positive effect of incentive measures on increasing vaccine uptake. Our model can be adapted to various national and regional contexts, making it a valuable tool for evaluating certain events. It also offers scalability to address future outbreaks, providing the government with valuable insights to effectively promote vaccines in response to different viral threats.

## Methods

### Data Source

We obtained daily reported COVID-19 infected cases (January 24, 2020, to October 15, 2022) and vaccination doses (February 26, 2021, to October 15, 2022) from the DATA.GOV.HK database^60^. According to the availability of vaccines, we divided the Hong Kong epidemic into three phases based on the vaccination plan: (1) pre-vaccination phase; from January 24, 2020, to February 25, 2021, during which the Hong Kong government relied solely on PHSMs to control the epidemic; (2) primary vaccination series phase; from February 26, 2021, to November 10, 2021, during which citizens could begin receiving the first two primary doses of Sinovac and BioNTech vaccines in addition to PHSMs; (3)booster vaccination phase; from November 11, 2021, to October 15, 2022, during which citizens could start receiving booster doses of the vaccine. We exclusively focused on locally acquired infections to isolate local transmission dynamics under Hong Kong’s stringent border quarantine policies. Five case categories - *epidemiologically linked with local case, epidemiologically linked with possibly local case, local case, locally acquired case, and possibly local case* – were consolidated into a unified *local transmission* category. A 7-day moving average was applied to mitigate reporting delays and weekday fluctuations in case counts. We aggregated Sinovac (CoronaVac) and BioNTech (Comirnaty) recipients for vaccination data, assuming comparable vaccine efficacy between the two vaccines. The contact matrix in Hong Kong was projected using empirical data from the COVID-19 era^61^. Additionally, daily mobility trends during the study period were extracted from Google community mobility reports^56^. We used the average changes in *retail and recreation percent change from baseline* and *transit stations percent change from baseline* to represent the extent of mobility changes in Hong Kong under different PHSMs implementations during the epidemic.

### Model Structure

We proposed a social-epidemiological transmission model based on the SEVPIRD-CT framework (susceptible, exposed, vaccinated, presymptomatic, infected, recovered, deceased, contact-traced), as illustrated in Supplementary, Fig. 1. Susceptible individuals enter an exposed (E) state upon infection, characterized by a latent period with no infectiousness. Following latency, they transition to a presymptomatic (P) state, becoming infectious but remaining asymptomatic. Presymptomatic individuals are either identified via contact tracing (CT) and isolated upon symptom onset or progress to the infected (I) state, with eventual transition to recovered (R) or deceased (D) compartments.

During the phase of vaccination programs, susceptible individuals move to a vaccinated (V) state upon immunization. Breakthrough infections in vaccinated individuals follow the same progression (𝐸 → 𝑃 → 𝐼 → 𝑅/𝐷) but with reduced mortality rates calibrated to vaccine efficacy. We incorporated imitation dynamics reflecting social learning processes to capture temporal shifts in vaccine uptake behavior. The daily vaccination willingness (𝑦^𝐻^) evolves according to:

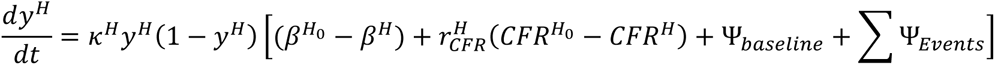

where 𝑦 represents the proportion of individuals vaccinated daily based on the daily maximum appointment vaccination capacity. 𝐻_0_ and 𝐻 act paired vaccination status (e.g., 𝐻_0_ = unvaccinated, 𝐻 = fully vaccinated; 𝐻_0_ = fully vaccinated, 𝐻 = received a booster dose). 𝜅𝜅 denotes the imitation rate at which individuals sample others and switch vaccinating behavior, while 𝛽 and 𝐶𝐹𝑅 stand for the transmission rate and case fatality rate of administering the dose 𝐻_0_ or 𝐻. 𝑟 is the adjustment coefficient for the difference in case fatality rates. Ψ_*Events*_ signifies the payoff gain following different vaccine-related events. The aggregated value of terms within the square brackets quantifies population-level risk perception, whereby the algebraic sign (positive/negative) governs the behavioral inclination toward vaccination—positive values amplify vaccine uptake through perceived benefit dominance, while negative values suppress it via risk aversion dominance.

### Statistical Analysis

We fitted the daily reported cases for each wave of the epidemic in Hong Kong, as well as the daily number of vaccinations during the vaccination program implementation period, using MCMC methods with the MCMCstat toolbox^64^. The model parameter values for each epidemic wave are presented in Supplementary Table 1. We randomly generated 10,000 sets of parameters using Latin Hypercube Sampling (LHS). After running all parameter sets, we selected the three sets with the most minor errors as our initial parameter sets.

For fitting each phase, we further divided each phase into segments based on the number of epidemic waves and the occurrence of certain events. For the first segment fitting, we assumed that an individual in the presymptomatic stage entered a population entirely susceptible to infection. The initial values for fitting each subsequent segment were set as the ending values of the previous segment. We used the Poisson likelihood function as the loss function for daily reported cases and vaccination numbers.

Finally, we estimated the convergence of the MCMC chains using the Gelman-Rubin diagnostic. Once the MCMC chains converged and stabilized, we randomly selected 500 parameter sets from the burn-in phase (half the chain length) to generate 95% confidence intervals based on these 500 parameter sets.

### Events Screening

A previous study surveyed Hong Kong to determine reasons for vaccine hesitancy across different age groups^49^. After excluding personal factors like concerns about chronic diseases, already antibodies, and lack of social norms or support, the top three reasons were identified for each group. These reasons were then grouped into vaccine-specific issues, low trust in the government, and a lack of urgency or perceived need for vaccination. To select relevant events, we applied polynomial fitting to daily vaccination data to find inflection points in the rate changes. These were identified on April 19, May 13, June 30, July 15, August 5, October 27, and November 7, 2021. Considering the lead or lag time of events^65^, related news was reviewed from the Department of Health press releases^66^ and the Oriental Daily News^67^ within 28 days before and after these dates. The events listed in Table 1 were then chosen, and their impact on vaccine hesitancy in Hong Kong was measured.

## Data availability

Data of COVID-19 reported cases, death numbers and count of vaccination were retrieved from official source (https://data.gov.hk/en-data/dataset/hk-dh-chpsebcddr-novel-infectious-agent (2023) and https://data.gov.hk/en-data/dataset/hk-hhb-hhbcovid19-vaccination-rates-over-time-by-age (2023)). Mobility changes were retrieved from (https://www.google.com/covid19/mobility/)

## Code availability

The fitting and analysis codes are publicly available at https://github.com/yifan7-chen/Vaccine_Hesitancy

## Data Availability

All data produced are available online at https://data.gov.hk/en/
https://www.google.com/covid19/mobility/
https://covid19.apple.com/mobility

https://www.google.com/covid19/mobility/

https://covid19.apple.com/mobility

## Acknowledgements

This work was supported in part by the Research Grants Council of the Hong Kong Special Administrative Region, China (Ref. No. 11218221, C7154-20GF, C7151-20GF and C1143-20GF to Q.Z.), and in part by the Health and Medical Research Fund (Ref. No: COVID190215 to H.-Y.Y. and Q.Z.). The funders had no role in study design, data collection and analysis, decision to publish or preparation of the manuscript.

## Author contributions

Q.Z. and Y.Y. designed and oversaw the study. Y.C. contributed to data collection, data analysis and data interpretation under the supervision of Q.Z. Y.C wrote the manuscript. Y.Y contributed to revising the manuscript. Q.Z. and H.Y.Y contributed to the interpretation of the findings and revising the manuscript. All authors approved the submitted version and have contributed to the final version of this manuscript.

