## Supplementary Information for "Impact assessment of vaccine-related negative news and incentive measures on vaccine hesitancy in Hong Kong"

Supplementary Methods

Individuals are divided into the following classes: susceptible individuals ($S$), individuals vaccinated with the first dose ($V_{1}$), individuals vaccinated with the second dose ($V_{2}$), individuals vaccinated with the booster dose ($V_{B}$), exposed individuals ($E$), presymptomatic individuals without vaccinal immunity ($P$), infectious individuals ($I$), contact-traced individuals ($CT$), recovered individuals ($R$), and deceased individuals ($D$). If these individuals have partial, full, or enhanced vaccinal immunity, we add a placeholder $V_{1}$, $V_{2}$, $V_{B}$ for each compartment, respectively. For example, exposed individuals with partial vaccinal immunity are presented by $E^{V_{1}}$.

We assume that vaccines are administrated in a 3-dose schedule, including a 2-dose primary series and a booster dose. Susceptible individuals are vaccinated with the first dose in the primary series at rate $y^{P}\left( t \right).$ We assume that no individuals who have received the first dose skip the second dose. The interval between the first and the second primary doses is $1/\omega$ days. Individuals who have received two doses in the primary series are vaccinated with the booster dose at the rate $y^{B}\left( t \right)$. We extend a social-epidemiological model with an imitation mechanism and payoff-driven risk perception to capture the evolution of vaccination behavior in the population^3,4^. Denote the payoffs to receive all doses in the primary series and the booster dose as $f_{v}^{P}$ and $f_{v}^{B}$, respectively.

$$f_{v}^{P}=\beta^{P}+r_{CFR}^{P}{CFR}^{P}$$

(1)

$$f_{v}^{B}=\beta^{B}+r_{CFR}^{B}{CFR}^{B}$$

(2)

The first term in (1)-(2) represents the perceived cost of being infected after vaccination. The second term in (1)-(2) models the perceived cost of death after vaccination.

The payoff not to receive all doses in the primary series ($f_{n}^{P}$) and the booster ($f_{n}^{B}$) are

$$f_{n}^{P}=\beta+r_{CFR}^{P}CFR$$

(3)

$$f_{n}^{B}=\beta^{P}+r_{CFR}^{B}{CFR}^{P}$$

(4)

which models the perceived cost of being infected and death without further vaccination.

The payoff of chosen events are

$$f_{Events}^{P}=\Psi_{baseline}^{P}+\sum\Psi_{Events}^{P}$$

(5)

$$f_{Events}^{B}=\Psi_{baseline}^{B}+\sum\Psi_{Events}^{B}$$

(6)

Then, the payoff gains for switching to receive all doses in the primary series and the booster dose are

$$\Delta E^{P}=f_{n}^{P}-f_{v}^{P}=\beta-\beta^{P}+r_{CFR}^{P}(CFR-{CFR}^{P})+\Psi_{baseline}^{P}+\sum\Psi_{Events}^{P}$$

(7)

and

$$\Delta E^{B}=f_{n}^{B}-f_{v}^{B}=\beta^{P}-\beta^{B}+r_{CFR}^{B}({CFR}^{P}-{CFR}^{B})+\Psi_{baseline}^{B}+\sum\Psi_{Events}^{B}$$

(8)

respectively.

According to the imitation mechanism,

$$\frac{dy^{P}}{dt}=\kappa^{P}y^{P}\left( 1-y^{P} \right)\left[ (\beta-\beta^{P})+r_{CFR}^{P}\left( CFR-{CFR}^{P} \right)+\Psi_{baseline}^{P}+\sum\Psi_{Events}^{P} \right]$$

(9)

$$\frac{dy^{B}}{dt}=\kappa^{B}y^{B}\left( 1-y^{B} \right)\left[ (\beta^{P}-\beta^{B})+r_{CFR}^{B}\left( {CFR}^{P}-{CFR}^{B} \right)+\Psi_{baseline}^{B}+\sum\Psi_{Events}^{B} \right]$$

(10)

The transition rates from $S$, $V_{1}$, $V_{2}$, and $V_{B}$ to $E$, $E^{V_{1}}$, $E^{V_{2}}$, and $E^{V_{B}}$ are $\lambda^{S}$, $\lambda^{V_{1}}$, $\lambda^{V_{2}}$, and $\lambda^{V_{B}}$, respectively.

$$\lambda^{S}\left( t \right)=\beta(t)\theta C(1-\varphi\left( t \right))\frac{\gamma^{Pre}({P\left( t \right)+P}^{V_{1}}\left( t \right)+P^{V_{2}}\left( t \right)+P^{V_{B}}(t))+\gamma({I\left( t \right)+I}^{V_{1}}\left( t \right)+I^{V_{2}}\left( t \right)+I^{V_{B}}(t))}{N}$$

(11)

$$\lambda^{V_{1}}\left( t \right)=(1-\eta^{1}(t))\beta(t)\theta C(1-\varphi\left( t \right))\frac{\gamma^{Pre}({P\left( t \right)+P}^{V_{1}}\left( t \right)+P^{V_{2}}\left( t \right)+P^{V_{B}}(t))+\gamma({I\left( t \right)+I}^{V_{1}}\left( t \right)+I^{V_{2}}\left( t \right)+I^{V_{B}}(t))}{N}$$

(12)

$$\lambda^{V_{2}}\left( t \right)=(1-\eta^{2}(t))\beta(t)\theta C(1-\varphi\left( t \right))\frac{\gamma^{Pre}({P\left( t \right)+P}^{V_{1}}\left( t \right)+P^{V_{2}}\left( t \right)+P^{V_{B}}(t))+\gamma({I\left( t \right)+I}^{V_{1}}\left( t \right)+I^{V_{2}}\left( t \right)+I^{V_{B}}(t))}{N}$$

(13)

$$\lambda^{V_{B}}\left( t \right)=(1-\eta^{B}(t))\beta(t)\theta C(1-\varphi\left( t \right))\frac{\gamma^{Pre}({P\left( t \right)+P}^{V_{1}}\left( t \right)+P^{V_{2}}\left( t \right)+P^{V_{B}}(t))+\gamma({I\left( t \right)+I}^{V_{1}}\left( t \right)+I^{V_{2}}\left( t \right)+I^{V_{B}}(t))}{N}$$

(14)

Here, $\beta$ is the transmissibility of combined strains. $\theta$ is the relative susceptibility to infection. $C$ is the average number of contacts among all age groups. $\varphi(t)$ is the effectiveness of public health and social measures (PHSMs), which are estimated by mobility changes. $\eta^{1}$, $\eta^{2}$, and $\eta^{B}$ are the 1-dose, 2-dose, booster-dose efficacy of vaccines against infection from combined strains.

Exposed individuals become infectious with a transition rate $\mu$. Infectious individuals become either recovered or deceased at a transition rate $\gamma$. Denote the severity of combined strains as $\epsilon(t)$. Denote the relative risk of mortality as $\xi$. Denote the 1-dose, 2-dose, and booster-dose efficacy of vaccines against death from combined strain as $\delta^{1}(t)$, $\delta^{2}(t)$, and $\delta^{B}(t)$, respectively. Thus, the transition rates from $I$, $I^{V_{1}}$, $I^{V_{2}}$, $I^{V_{B}}$ to $R$ are $\gamma\left( 1-\epsilon\left( t \right)\xi\right)$, $\gamma\left[ 1-\epsilon(t)\left( 1-\delta^{1} \right)\xi\right]$, $\gamma\left[ 1-\epsilon\left( t \right)\left( 1-\delta^{2} \right)\xi\right]$, $\gamma\left[ 1-\epsilon\left( t \right)\left( 1-\delta^{B} \right)\xi\right]$, respectively. The transition rates from $I$, $I^{V_{1}}$, $I^{V_{2}}$, $I^{V_{B}}$ to $R$ are $\gamma\epsilon\left( t \right)\xi$, $\gamma\epsilon\left( t \right)\left( 1-\delta^{1} \right)\xi$, $\gamma\epsilon\left( t \right)\left( 1-\delta^{2} \right)\xi$, $\gamma\epsilon\left( t \right)\left( 1-\delta^{B} \right)\xi$, respectively.

Model Equations

Phase 1:

$$\dot{S}\left( t \right)= -\lambda^{S}\left( t \right)S\left( t \right)$$

$$\dot{E}\left( t \right)= \lambda^{S}\left( t \right)S\left( t \right)-\mu E(t)$$

$$\dot{P}\left( t \right)= \mu E\left( t \right)-D_{p}P(t)$$

$$\dot{I}\left( t \right)= D_{p}P\left( t \right)-p_{CT}I\left( t \right)-\gamma I(t)$$

$$\dot{CT}\left( t \right)= p_{CT}I\left( t \right)- \omega_{CT}CT(t)$$

$$\dot{R}\left( t \right)=\left( 1-\epsilon\left( t \right)\xi\right)[\gamma I\left( t \right)+ \omega_{CT}CT\left( t \right)]$$

$$\dot{D}\left( t \right)=\epsilon\left( t \right)\xi[\gamma I\left( t \right)+ \omega_{CT}CT\left( t \right)]$$

(15)

Phase 2:

|  | $\dot{S}\left( t \right)= -\lambda^{S}\left( t \right)S\left( t \right)-\Phi y^{P}\left( t \right)$ $\dot{V}^{1}\left( t \right)=\Phi y^{P}\left( t \right)-\lambda^{V_{1}}\left( t \right)V^{1}\left( t \right)-\omega V^{1}\left( t \right)$ $\dot{V}^{2}\left( t \right)=-\lambda^{V_{2}}\left( t \right)V^{2}\left( t \right)+\omega V^{1}\left( t \right)$ $\dot{E}\left( t \right)= \lambda^{S}\left( t \right)S\left( t \right)-\mu E(t)$ $\dot{E}^{V_{1}}\left( t \right)= \lambda^{V_{1}}\left( t \right)V^{1}\left( t \right)-\mu E^{V_{1}}(t)$ $\dot{E}^{V_{2}}\left( t \right)= \lambda^{V_{2}}\left( t \right)V^{2}\left( t \right)-\mu E^{V_{2}}(t)$ $\dot{P}\left( t \right)= \mu E\left( t \right)-D_{p}P(t)$ $\dot{P}^{V_{1}}\left( t \right)= \mu E^{V_{1}}\left( t \right)-D_{p}P^{V_{1}}\left( t \right)$ $\dot{P}^{V_{2}}\left( t \right)= \mu E^{V_{2}}\left( t \right)-D_{p}P^{V_{2}}(t)$ $\dot{I}\left( t \right)= D_{p}P\left( t \right)-p_{CT}I\left( t \right)-\gamma I(t)$ $\dot{I}^{V_{1}}\left( t \right)= D_{p}P^{V_{1}}\left( t \right)-p_{CT}I^{V_{1}}\left( t \right)-\gamma I^{V_{1}}(t)$ $\dot{I}^{V_{2}}\left( t \right)= D_{p}P^{V_{2}}\left( t \right)-p_{CT}I^{V_{2}}\left( t \right)-\gamma I^{V_{2}}(t)$ $\dot{CT}\left( t \right)= p_{CT}I\left( t \right)- \omega_{CT}CT(t)$ $\dot{CT}^{V_{1}}\left( t \right)= p_{CT}I^{V_{1}}\left( t \right)- \omega_{CT}{CT}^{V_{1}}(t)$ $\dot{CT}^{V_{2}}\left( t \right)= p_{CT}I^{V_{2}}\left( t \right)- \omega_{CT}{CT}^{V_{2}}(t)$  $\dot{R}\left( t \right)=\left( 1-\epsilon\left( t \right)\xi\right)[\gamma I\left( t \right)+ \omega_{CT}CT\left( t \right)]$ *+*$\left( 1-\epsilon\left( t \right)(1-\delta^{1})\xi\right)[\gamma I^{V_{1}}\left( t \right)+ \omega_{CT}{CT}^{V_{1}}\left( t \right)]$ *+* $\left( 1-\epsilon\left( t \right)(1-\delta^{2})\xi\right)[\gamma I^{V_{2}}\left( t \right)+ \omega_{CT}{CT}^{V_{2}}\left( t \right)]$  $\dot{D}\left( t \right)=\epsilon\left( t \right)\xi[\gamma I\left( t \right)+ \omega_{CT}CT\left( t \right)]$ *+*$\epsilon\left( t \right)(1-\delta^{1})\xi[\gamma I^{V_{1}}\left( t \right)+ \omega_{CT}{CT}^{V_{1}}\left( t \right)]$ *+* $\epsilon\left( t \right)(1-\delta^{2})\xi[\gamma I^{V_{2}}\left( t \right)+ \omega_{CT}{CT}^{V_{2}}\left( t \right)]$ | (16) |
| --- | --- | --- |

Phase 3:

|  | $\dot{S}\left( t \right)= -\lambda^{S}\left( t \right)S\left( t \right)-\Phi y^{P}\left( t \right)$ $\dot{V}^{1}\left( t \right)=\Phi y^{P}\left( t \right)-\lambda^{V_{1}}\left( t \right)V^{1}\left( t \right)-\omega V^{1}\left( t \right)$ $\dot{V}^{2}\left( t \right)=-\lambda^{V_{2}}\left( t \right)V^{2}\left( t \right)+\omega V^{1}\left( t \right)$ $\dot{V}^{B}\left( t \right)=\Phi y^{B}\left( t \right)-\lambda^{V_{B}}\left( t \right)V^{B}\left( t \right)$ $\dot{E}\left( t \right)= \lambda^{S}\left( t \right)S\left( t \right)-\mu E(t)$ $\dot{E}^{V_{1}}\left( t \right)= \lambda^{V_{1}}\left( t \right)V^{1}\left( t \right)-\mu E^{V_{1}}(t)$ $\dot{E}^{V_{2}}\left( t \right)= \lambda^{V_{2}}\left( t \right)V^{2}\left( t \right)-\mu E^{V_{2}}(t)$ $\dot{E}^{V_{B}}\left( t \right)= \lambda^{V_{B}}\left( t \right)V^{B}\left( t \right)-\mu E^{V_{B}}(t)$ $\dot{P}\left( t \right)= \mu E\left( t \right)-D_{p}P(t)$ $\dot{P}^{V_{1}}\left( t \right)= \mu E^{V_{1}}\left( t \right)-D_{p}P^{V_{1}}\left( t \right)$ $\dot{P}^{V_{2}}\left( t \right)= \mu E^{V_{2}}\left( t \right)-D_{p}P^{V_{2}}(t)$ $\dot{P}^{V_{B}}\left( t \right)= \mu E^{V_{B}}\left( t \right)-D_{p}P^{V_{B}}(t)$ $\dot{I}\left( t \right)= D_{p}P\left( t \right)-p_{CT}I\left( t \right)-\gamma I(t)$ $\dot{I}^{V_{1}}\left( t \right)= D_{p}P^{V_{1}}\left( t \right)-p_{CT}I^{V_{1}}\left( t \right)-\gamma I^{V_{1}}(t)$ $\dot{I}^{V_{2}}\left( t \right)= D_{p}P^{V_{2}}\left( t \right)-p_{CT}I^{V_{2}}\left( t \right)-\gamma I^{V_{2}}(t)$ $\dot{I}^{V_{B}}\left( t \right)= D_{p}P^{V_{B}}\left( t \right)-p_{CT}I^{V_{B}}\left( t \right)-\gamma I^{V_{B}}(t)$ $\dot{CT}\left( t \right)= p_{CT}I\left( t \right)- \omega_{CT}CT(t)$ $\dot{CT}^{V_{1}}\left( t \right)= p_{CT}I^{V_{1}}\left( t \right)- \omega_{CT}{CT}^{V_{1}}(t)$ $\dot{CT}^{V_{2}}\left( t \right)= p_{CT}I^{V_{2}}\left( t \right)- \omega_{CT}{CT}^{V_{2}}(t)$ $\dot{CT}^{V_{B}}\left( t \right)= p_{CT}I^{V_{B}}\left( t \right)- \omega_{CT}{CT}^{V_{B}}(t)$  $\dot{R}\left( t \right)=\left( 1-\epsilon\left( t \right)\xi\right)[\gamma I\left( t \right)+ \omega_{CT}CT\left( t \right)]$ *+*$\left( 1-\epsilon\left( t \right)(1-\delta^{1})\xi\right)[\gamma I^{V_{1}}\left( t \right)+ \omega_{CT}{CT}^{V_{1}}\left( t \right)]$ *+* $\left( 1-\epsilon\left( t \right)(1-\delta^{2})\xi\right)\left[ \gamma I^{V_{2}}\left( t \right)+ \omega_{CT}{CT}^{V_{2}}\left( t \right) \right]+ \left( 1-\epsilon\left( t \right)(1-\delta^{B})\xi\right)[\gamma I^{V_{B}}\left( t \right)+ \omega_{CT}{CT}^{V_{B}}\left( t \right)]$  $\dot{D}\left( t \right)=\epsilon\left( t \right)\xi[\gamma I\left( t \right)+ \omega_{CT}CT\left( t \right)]$ *+*$\epsilon\left( t \right)(1-\delta^{1})\xi[\gamma I^{V_{1}}\left( t \right)+ \omega_{CT}{CT}^{V_{1}}\left( t \right)]$ *+* $\epsilon\left( t \right)\left( 1-\delta^{2} \right)\xi\left[ \gamma I^{V_{2}}\left( t \right)+ \omega_{CT}{CT}^{V_{2}}\left( t \right) \right]+\epsilon\left( t \right)(1-\delta^{B})\xi[\gamma I^{V_{B}}\left( t \right)+ \omega_{CT}{CT}^{V_{B}}\left( t \right)]$ | (17) |
| --- | --- | --- |

Supplementary Table 1: Parameters definitions, values and sources

| **Parameter** | **Meaning** | **Value** |
| --- | --- | --- |
| $\boldsymbol{N}_{\boldsymbol{a}}$ | The population size in Hong Kong | 0-4: 228994; 5-9: 289446;  10-14: 289398; 15-19: 264900;  20-24: 326214; 25-29: 458407;  30-34: 527028; 35-39: 587743;  40-44: 586611; 45-49: 582989;  50-54: 575242; 55-59: 630782;  60-64: 613802; 65-69: 492235;  70-74: 370751; 75+: 588528^5^ |
| $\boldsymbol{\kappa}^{\boldsymbol{P}}$ | Social learning rate of primary doses vaccination | Fitted |
| $\boldsymbol{\kappa}^{\boldsymbol{B}}$ | Social learning rate of booster doses vaccination | Fitted |
| $\boldsymbol{r}^{\boldsymbol{P}}$ | The adjustment coefficient for the difference in death rates after primary doses vaccination | Fitted |
| $\boldsymbol{r}^{\boldsymbol{B}}$ | The adjustment coefficient for the difference in death rates after booster dose vaccination | Fitted |
| $\boldsymbol{\Psi}_{\boldsymbol{Baseline}}$ | The baseline payoff gain under different contexts | Fitted |
| $\boldsymbol{\Psi}_{\boldsymbol{Events}}$ | The payoff gain following different events, such as vaccine packaging issues, government incentive policies, etc | Fitted |
| $\boldsymbol{\Phi}$ | The maximum supply of doses each day | 50000^6^ |
| $\boldsymbol{\beta}_{\boldsymbol{i}}$ | The transmissibility of strain i | Original strain: $\beta_{0}=0.558$; Alpha: 1.29$\beta_{0}$; Beta: 1.25$\beta_{0}$; Delta: 1.97$\beta_{0}$; Omicron: 1.97${\times4.31\beta}_{0} ADDIN ZOTERO\_ITEM CSL\_CITATION \{"citationID":"WSUZwuny","properties":\{"formattedCitation":"\backslash\backslash super 7,8\backslash\backslash nosupersub\{\}","plainCitation":"7,8","noteIndex":0\},"citationItems":[\{"id":1575,"uris":["http://zotero.org/users/8960282/items/IXRC89KT"],"itemData":\{"id":1575,"type":"article-journal","abstract":"We present a global analysis of the spread of recently emerged SARS-CoV-2 variants and estimate changes in effective reproduction numbers at country-specific level using sequence data from GISAID. Nearly all investigated countries demonstrated rapid replacement of previously circulating lineages by the World Health Organization-designated variants of concern, with estimated transmissibility increases of 29\% (95\% CI: 24-33), 25\% (95\% CI: 20-30), 38\% (95\% CI: 29-48) and 97\% (95\% CI: 76-117), respectively, for B.1.1.7, B.1.351, P.1 and B.1.617.2.","archive\_location":"410 citation(s)","call-number":"21.286","container-title":"Eurosurveillance","DOI":"10.2807/1560-7917.ES.2021.26.24.2100509","ISSN":"1560-7917","issue":"24","language":"en","note":"TLDR: A global analysis of the spread of recently emerged SARS-CoV-2 variants and estimate changes in effective reproduction numbers at country-specific level using sequence data from GISAID shows rapid replacement of previously circulating lineages by the World Health Organization-designated variants of concern.","source":"2","title":"Increased transmissibility and global spread of SARS-CoV-2 variants of concern as at June 2021","URL":"https://www.eurosurveillance.org/content/10.2807/1560-7917.ES.2021.26.24.2100509","volume":"26","author":[\{"family":"Campbell","given":"Finlay"\},\{"family":"Archer","given":"Brett"\},\{"family":"Laurenson-Schafer","given":"Henry"\},\{"family":"Jinnai","given":"Yuka"\},\{"family":"Konings","given":"Franck"\},\{"family":"Batra","given":"Neale"\},\{"family":"Pavlin","given":"Boris"\},\{"family":"Vandemaele","given":"Katelijn"\},\{"family":"Van Kerkhove","given":"Maria D"\},\{"family":"Jombart","given":"Thibaut"\},\{"family":"Morgan","given":"Oliver"\},\{"family":"Polain de Waroux","given":"Olivier","non-dropping-particle":"le"\}],"accessed":\{"date-parts":[["2022",5,31]]\},"issued":\{"date-parts":[["2021",6,17]]\}\}\},\{"id":1574,"uris":["http://zotero.org/users/8960282/items/ASND2CY2"],"itemData":\{"id":1574,"type":"article-journal","archive\_location":"4 citation(s)","call-number":"38.104","container-title":"Signal Transduction and Targeted Therapy","DOI":"10.1038/s41392-022-01009-8","ISSN":"2059-3635","issue":"1","journalAbbreviation":"Sig Transduct Target Ther","language":"en","note":"TLDR: Analysis using advanced structure determining methods such as cryo-electron microscopy and X-ray crystallography results directly connected novel mutations and new chemical interaction sites.","page":"151","source":"1","title":"Omicron: increased transmissibility and decreased pathogenicity","title-short":"Omicron","volume":"7","author":[\{"family":"Bálint","given":"Gábor"\},\{"family":"Vörös-Horváth","given":"Barbara"\},\{"family":"Széchenyi","given":"Aleksandar"\}],"issued":\{"date-parts":[["2022",12]]\}\}\}],"schema":"https://github.com/citation-style-language/schema/raw/master/csl-citation.json"\}$^7,8^ |
| $\boldsymbol{\theta}_{\boldsymbol{a}}$ | The relative susceptibility to infection for age group a | 0-9: 0.33; 10-19: 0.37;  20-29: 0.69; 30-39: 0.81;  40-49: 0.74; 50-59: 0.8;  60-69: 0.89; 70-79: 0.77;  80+: 0.77^9^ |
| $\boldsymbol{\eta}_{\boldsymbol{i}}^{\boldsymbol{1}}$ | The 1-dose efficacy of vaccines against infection from strain i | Original strain: 0.63; Alpha: 0.67;  Beta: 0.50; Delta: 0.57^10^;  Omicron: 0^11^ |
| $\boldsymbol{\eta}_{\boldsymbol{i}}^{\boldsymbol{2}}$ | The 2-dose efficacy of vaccines against infection from strain i | Original strain: 0.92; Alpha: 0.88;  Beta: 0.86^10^; Delta: 0.89;  Omicron: 0.36^12^ |
| $\boldsymbol{\eta}_{\boldsymbol{i}}^{\boldsymbol{B}}$ | The booster-dose efficacy of vaccines against infection from strain i | Delta: 0.97; Omicron: 0.6^12^ |
| $\boldsymbol{\mu}$ | The mean latent period | 2.9^13^ |
| $\boldsymbol{D}_{\boldsymbol{p}}$ | The mean presymptomatic infectious period | 2.3^13^ |
| $\boldsymbol{\gamma}$ | The mean symptomatic infectious period | 2.9^13^ |
| $\boldsymbol{p}_{\boldsymbol{CT}}$ | The probability of being contact traced | 0.2273 (Estimated) |

| Supplementary Table 2. Contact Matrix in Hong Kong^14^ | 75+ | 0.05 | 0.04 | 0.05 | 0.02 | 0.05 | 0.03 | 0.07 | 0.06 | 0.07 | 0.12 | 0.11 | 0.13 | 0.17 | 0.19 | 0.35 | 0.22 |
| --- | --- | --- | --- | --- | --- | --- | --- | --- | --- | --- | --- | --- | --- | --- | --- | --- | --- |
|  | 70-74 | 0.1 | 0.05 | 0.06 | 0.03 | 0.06 | 0.05 | 0.07 | 0.15 | 0.14 | 0.13 | 0.13 | 0.18 | 0.4 | 0.39 | 0.93 | 0.25 |
|  | 65-69 | 0.14 | 0.1 | 0.08 | 0.05 | 0.06 | 0.09 | 0.15 | 0.25 | 0.2 | 0.15 | 0.18 | 0.36 | 0.65 | 1.04 | 0.75 | 0.28 |
|  | 60-64 | 0.18 | 0.15 | 0.08 | 0.06 | 0.16 | 0.27 | 0.37 | 0.44 | 0.38 | 0.31 | 0.47 | 0.84 | 1.44 | 0.76 | 0.82 | 0.2 |
|  | 55-59 | 0.24 | 0.15 | 0.1 | 0.1 | 0.34 | 0.56 | 0.7 | 0.62 | 0.51 | 0.62 | 1.07 | 1.77 | 1.25 | 0.73 | 0.46 | 0.25 |
|  | 50-54 | 0.28 | 0.14 | 0.17 | 0.24 | 0.63 | 1.04 | 0.95 | 1.05 | 1.24 | 1.27 | 1.97 | 1.59 | 1.06 | 0.66 | 0.58 | 0.39 |
|  | 45-49 | 0.24 | 0.23 | 0.33 | 0.46 | 0.88 | 1.07 | 1.19 | 1.39 | 1.75 | 2.15 | 1..91 | 1.26 | 1.06 | 0.57 | 0.71 | 0.39 |
|  | 40-44 | 0.41 | 0.52 | 0.56 | 0.45 | 0.74 | 1.28 | 1.48 | 2.11 | 2.88881 | 1.64 | 1.73 | 1.41 | 1.25 | 0.98 | 0.92 | 0.37 |
|  | 35-39 | 0.72 | 0.59 | 0.42 | 0.34 | 0.84 | 1.52 | 1.96 | 3.04 | 1.96 | 1.52 | 1.32 | 1.32 | 1.49 | 1.05 | 0.87 | 0.37 |
|  | 30-34 | 0.78 | 0.48 | 0.26 | 0.23 | 0.97 | 1.97 | 2.91 | 1.89 | 1.75 | 1.33 | 1.40 | 1.76 | 1.46 | 1.09 | 0.5 | 0.28 |
|  | 25-29 | 0.55 | 0.3 | 0.17 | 0.32 | 1.47 | 3.45 | 1.91 | 1.52 | 1.35 | 1.14 | 1.57 | 1.65 | 1.27 | 0.76 | 0.54 | 0.17 |
|  | 20-24 | 0.34 | 0.15 | 0.22 | 0.71 | 3.05 | 1.79 | 0.95 | 0.69 | 0.88 | 0.95 | 1.15 | 1.01 | 0.79 | 0.57 | 0.28 | 0.17 |
|  | 15-19 | 0.26 | 0.23 | 0.70 | 9.53 | 2.11 | 0.50 | 0.28 | 0.51 | 0.85 | 1.13 | 0.96 | 0.87 | 0.77 | 0.54 | 0.83 | 0.44 |
|  | 10-14 | 0.40 | 0.98 | 10.92 | 3.23 | 0.24 | 0.15 | 0.53 | 0.95 | 1.22 | 0.82 | 0.78 | 0.69 | 0.77 | 0.99 | 0.94 | 0.55 |
|  | 5-9 | 0.97 | 8.08 | 2.15 | 0.28 | 0.2 | 0.28 | 0.8 | 1.25 | 1.00 | 0.64 | 0.57 | 1.07 | 1.25 | 1.29 | 1.22 | 0.41 |
|  | 0-4 | 2.00 | 0.83 | 0.19 | 0.12 | 0.24 | 0.63 | 0.84 | 000.92 | 0.57 | 0.34 | 0.56 | 1.07 | 1.35 | 0.92 | 0.47 | 0.33 |
|  | Contactee  Age Group  Contactor  Age Group | 0-4 | 5-9 | 10-14 | 15-19 | 20-24 | 25-29 | 30-34 | 35-39 | 40-44 | 45-49 | 50-54 | 55-59 | 60-64 | 65-69 | 70-74 | 75+ |

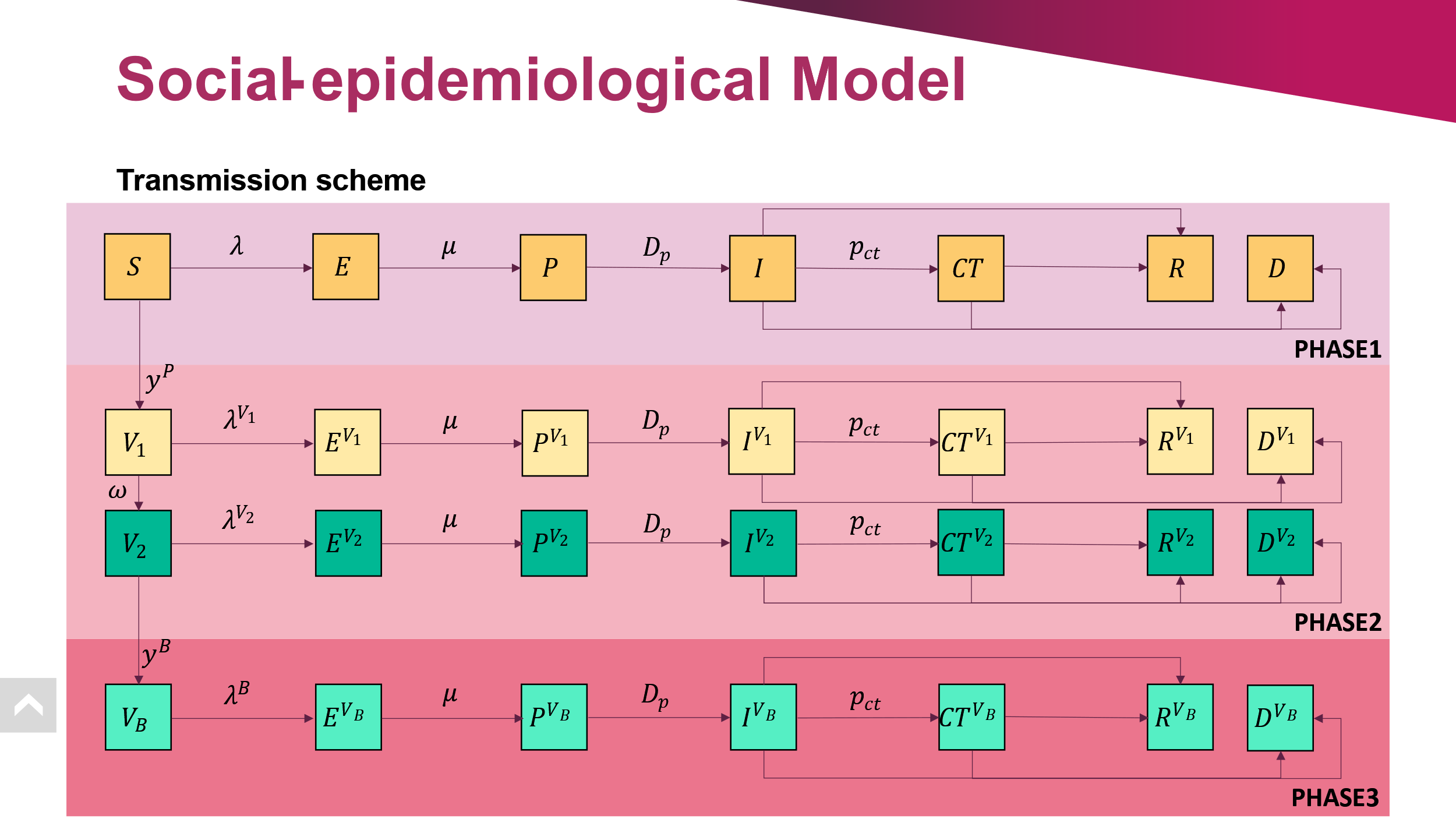

Supplementary Figure 1. Transmission scheme.

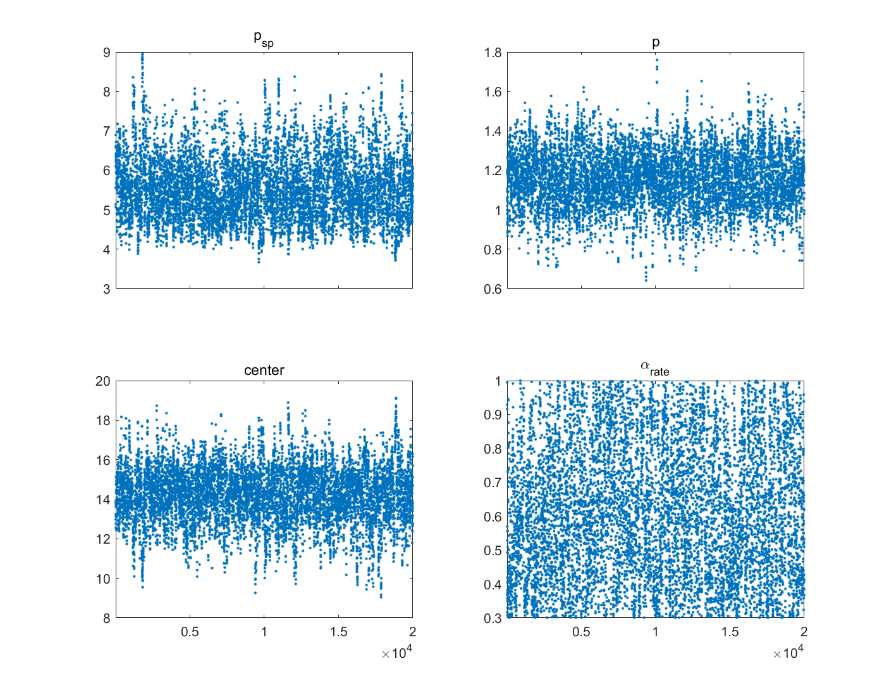

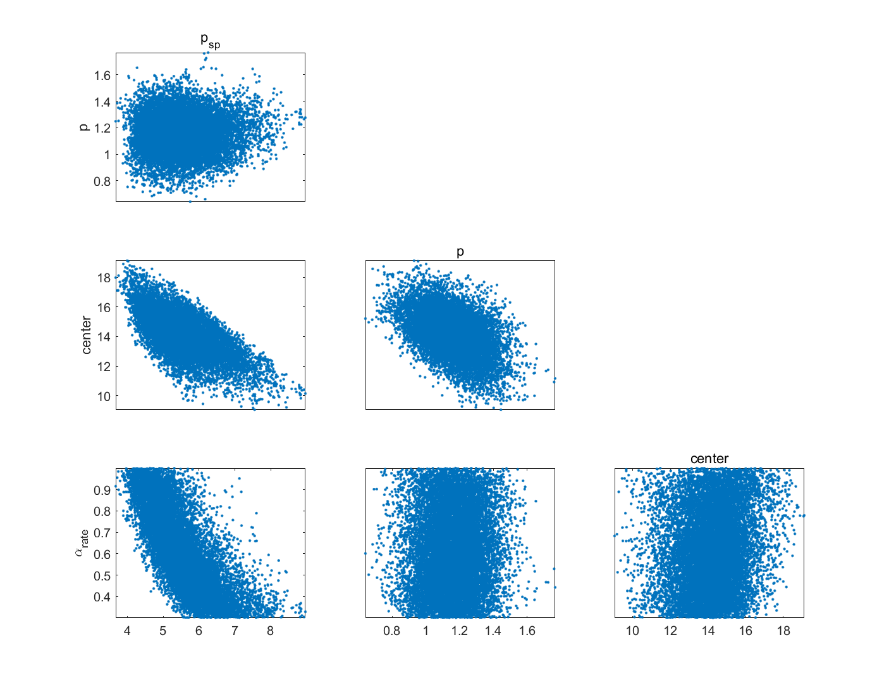

Supplementary Figure 2. Chain-panel plot and pairwise parameter correlations during the first segment (Jan. 24, 2020 – Mar. 02, 2020) in Phase 1.

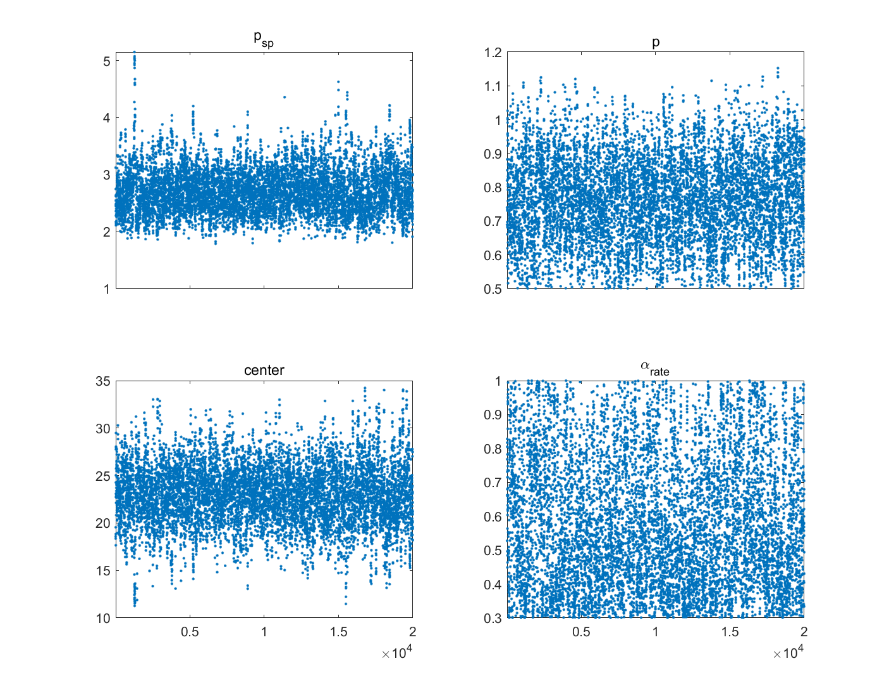

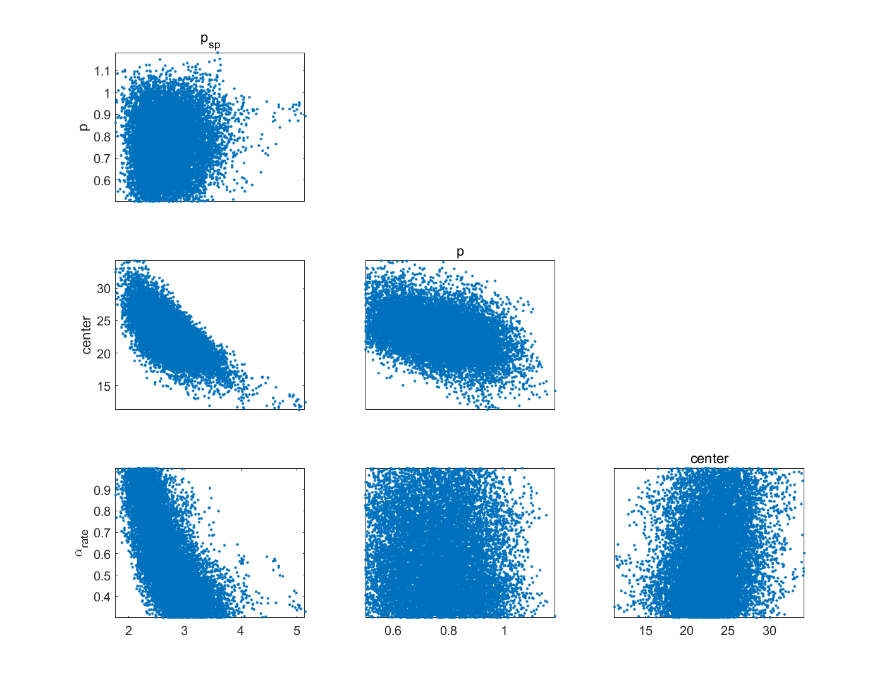

Supplementary Figure 3. Chain-panel plot and pairwise parameter correlations during the second segment (Mar. 02, 2020 – Jun. 28, 2020) in Phase 1.

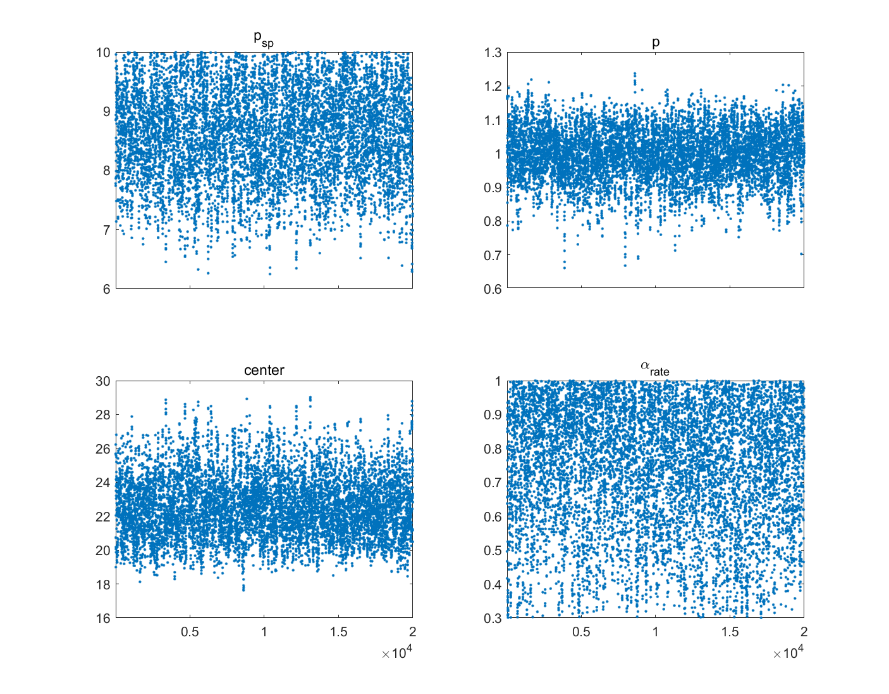

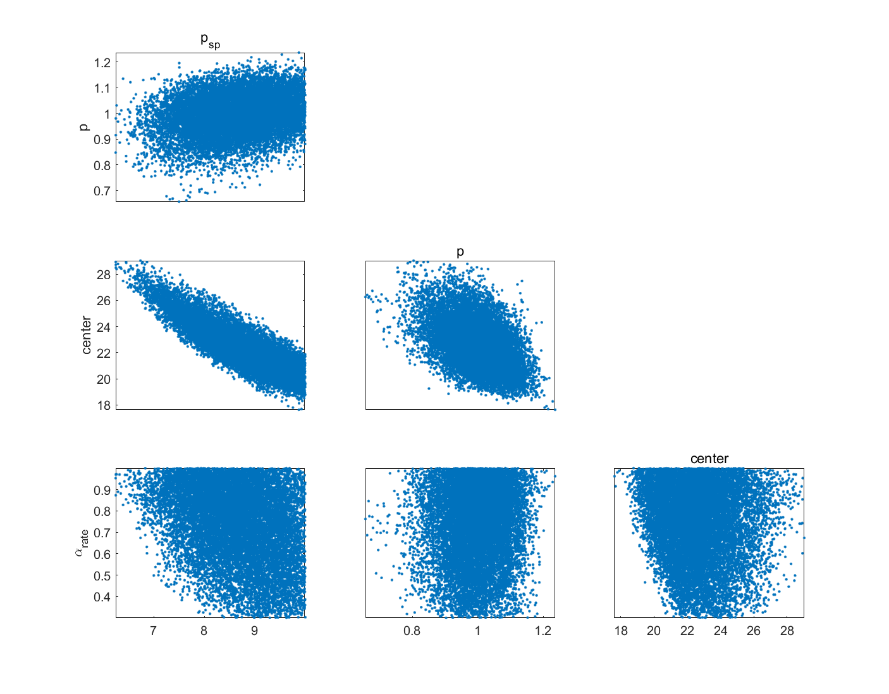

Supplementary Figure 4. Chain-panel plot and pairwise parameter correlations during the third segment (Jun. 28, 2020 – Oct. 28, 2020) in Phase 1.

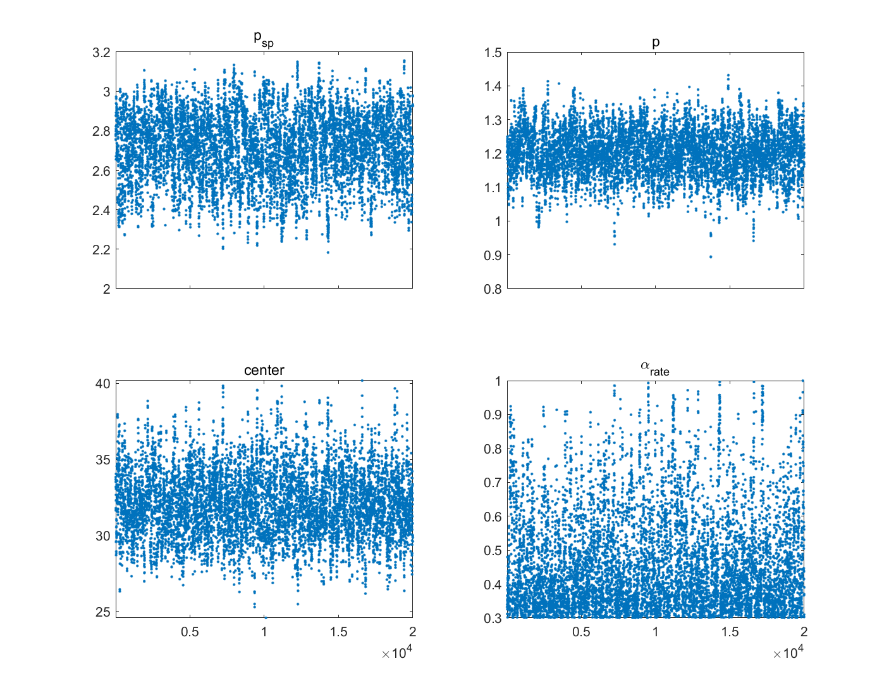

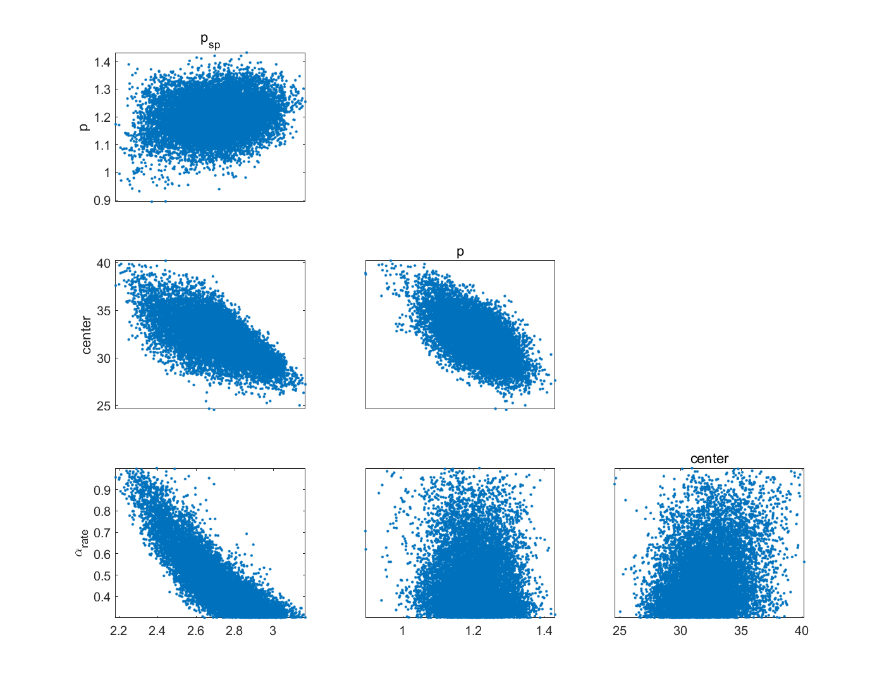

Supplementary Figure 5. Chain-panel plot and pairwise parameter correlations during the fourth segment (Oct. 28, 2020 – Jan. 04, 2021) in Phase 1.

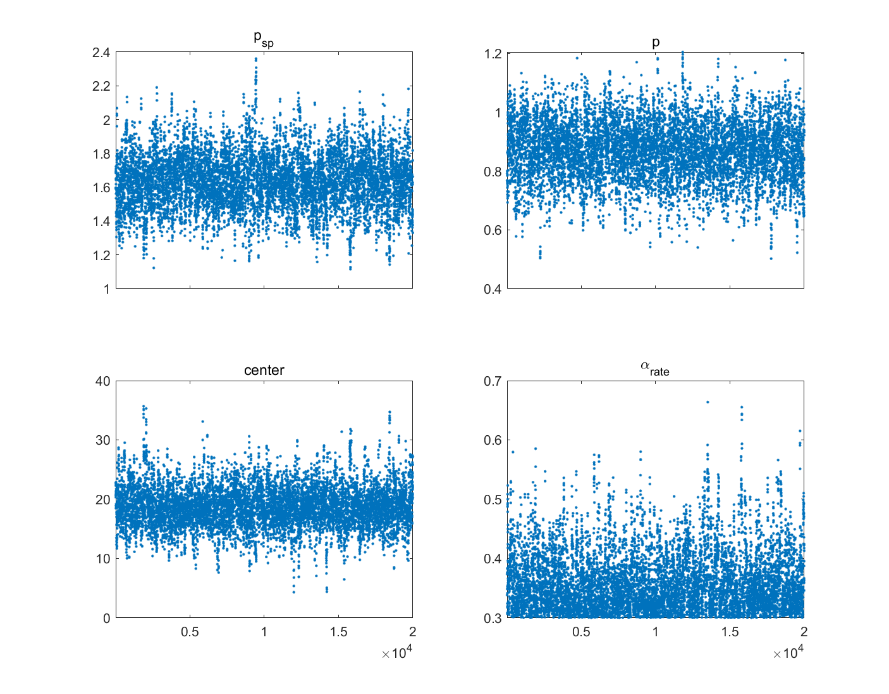

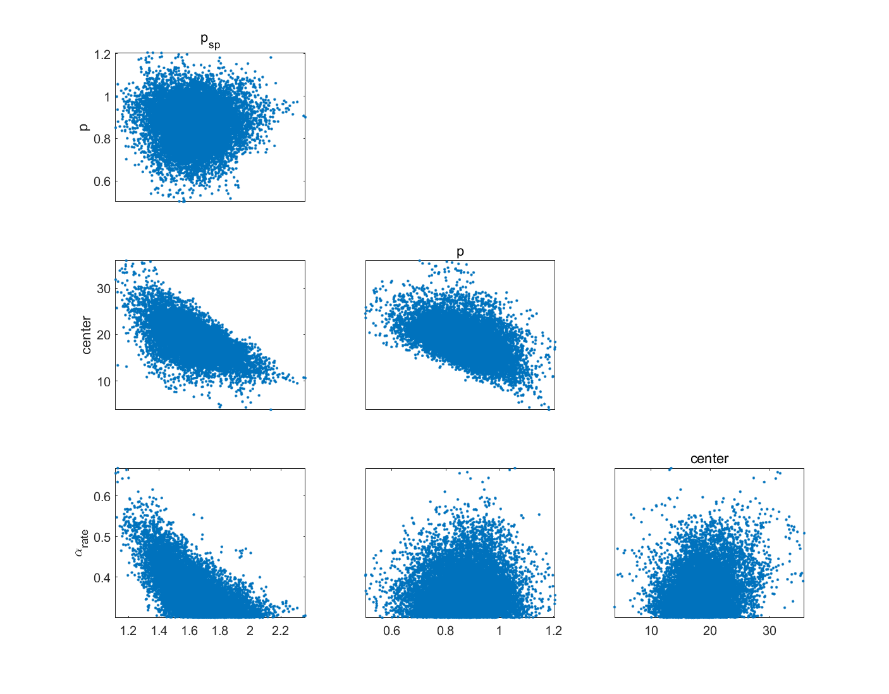

Supplementary Figure 6. Chain-panel plot and pairwise parameter correlations during the fifth segment (Jan. 04, 2021 – Feb. 26, 2021) in Phase 1.

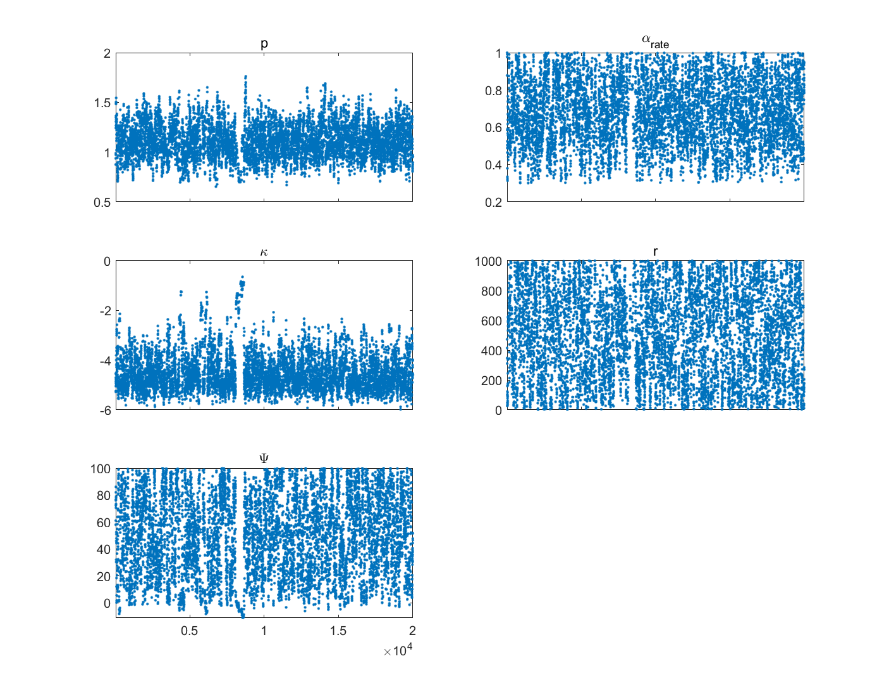

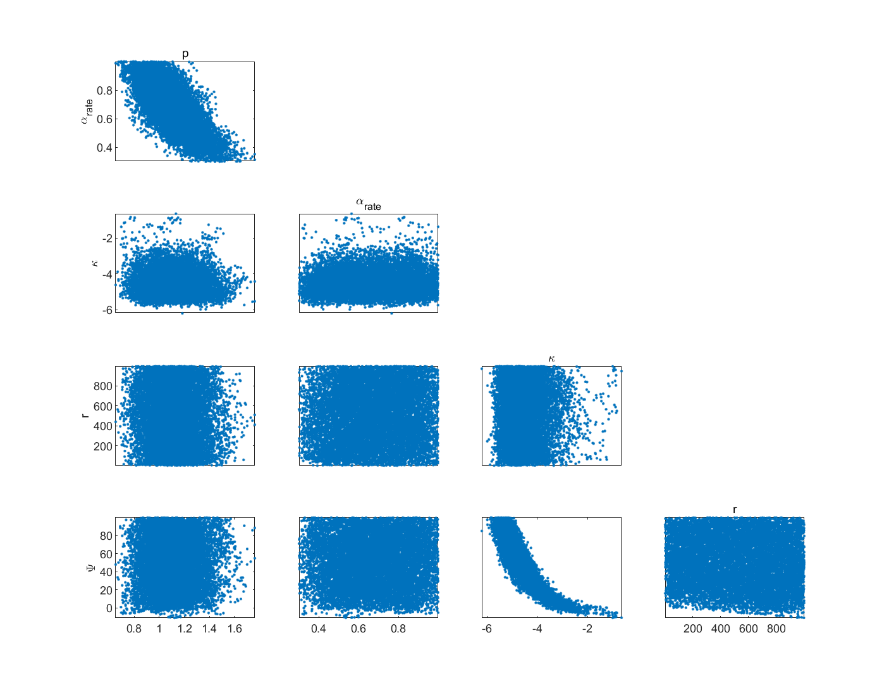

Supplementary Figure 7. Chain-panel plot and pairwise parameter correlations during the first segment (Feb. 26, 2021 – Mar. 23, 2021) in Phase 2.

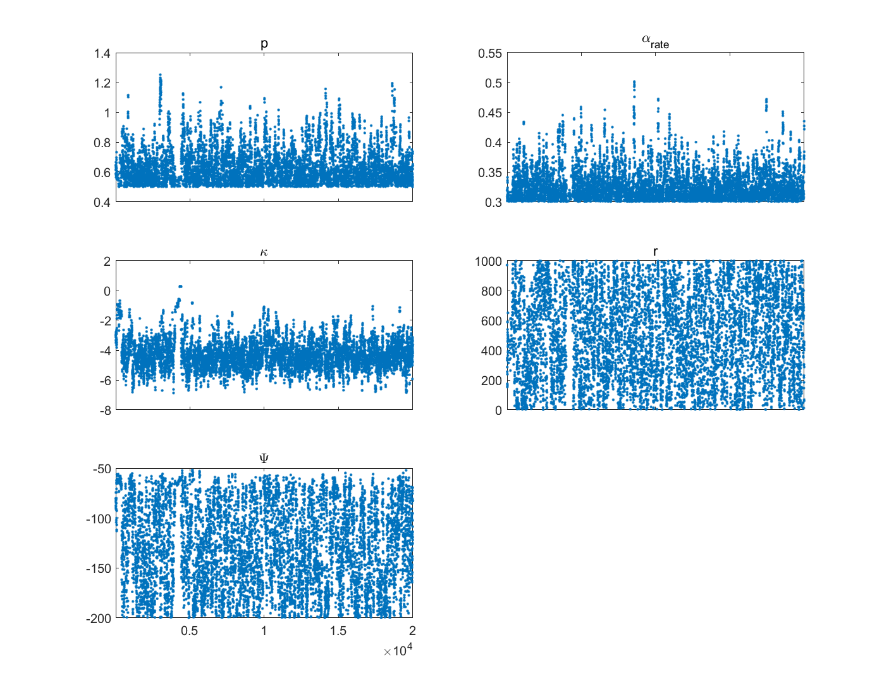

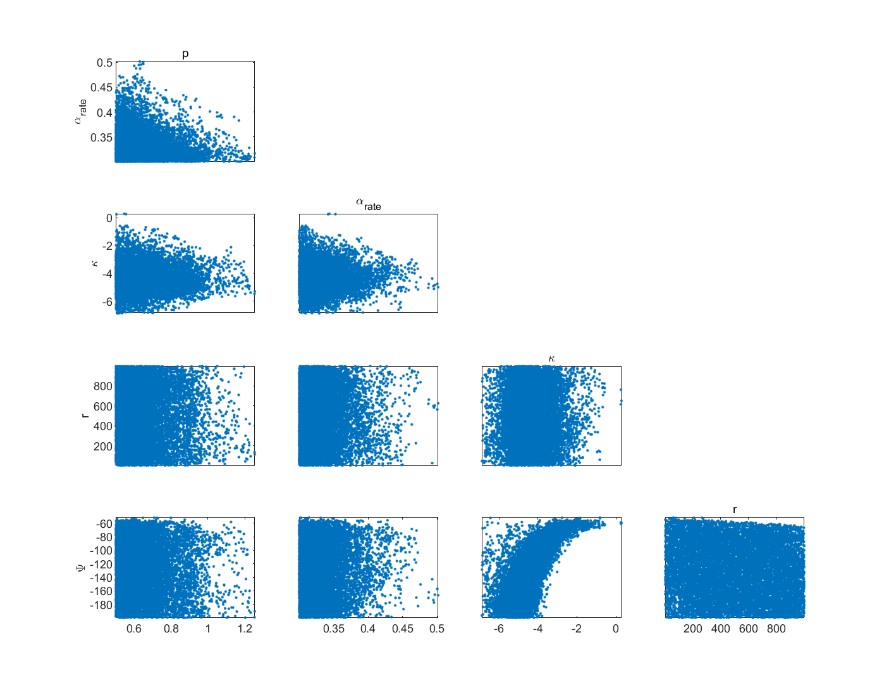

Supplementary Figure 8. Chain-panel plot and pairwise parameter correlations during the second segment (Mar. 24, 2021 – Apr. 04, 2021) in Phase 2.

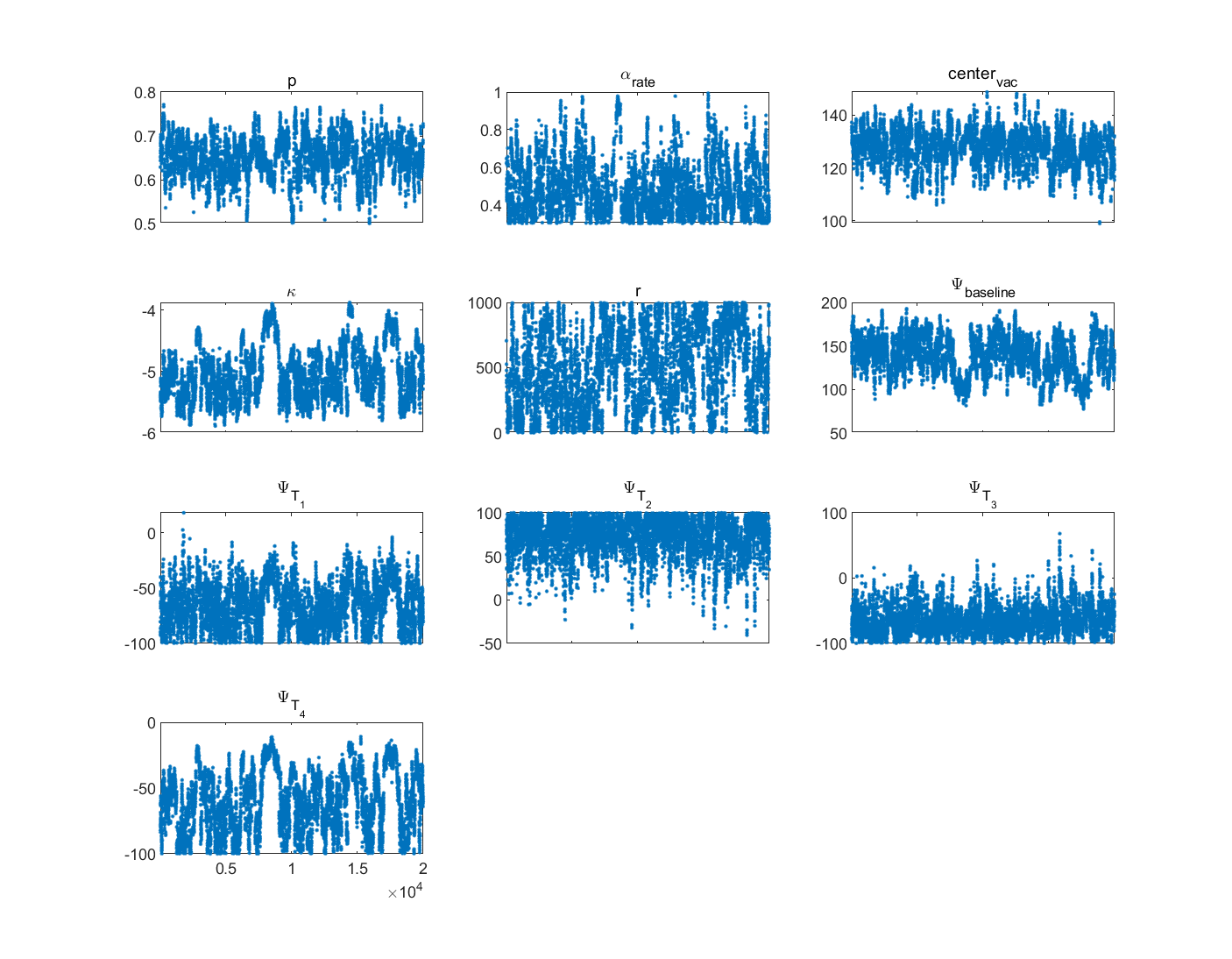

Supplementary Figure 9. Chain-panel plot during the third segment (Apr. 05, 2021 – Nov. 11, 2021) in Phase 2.

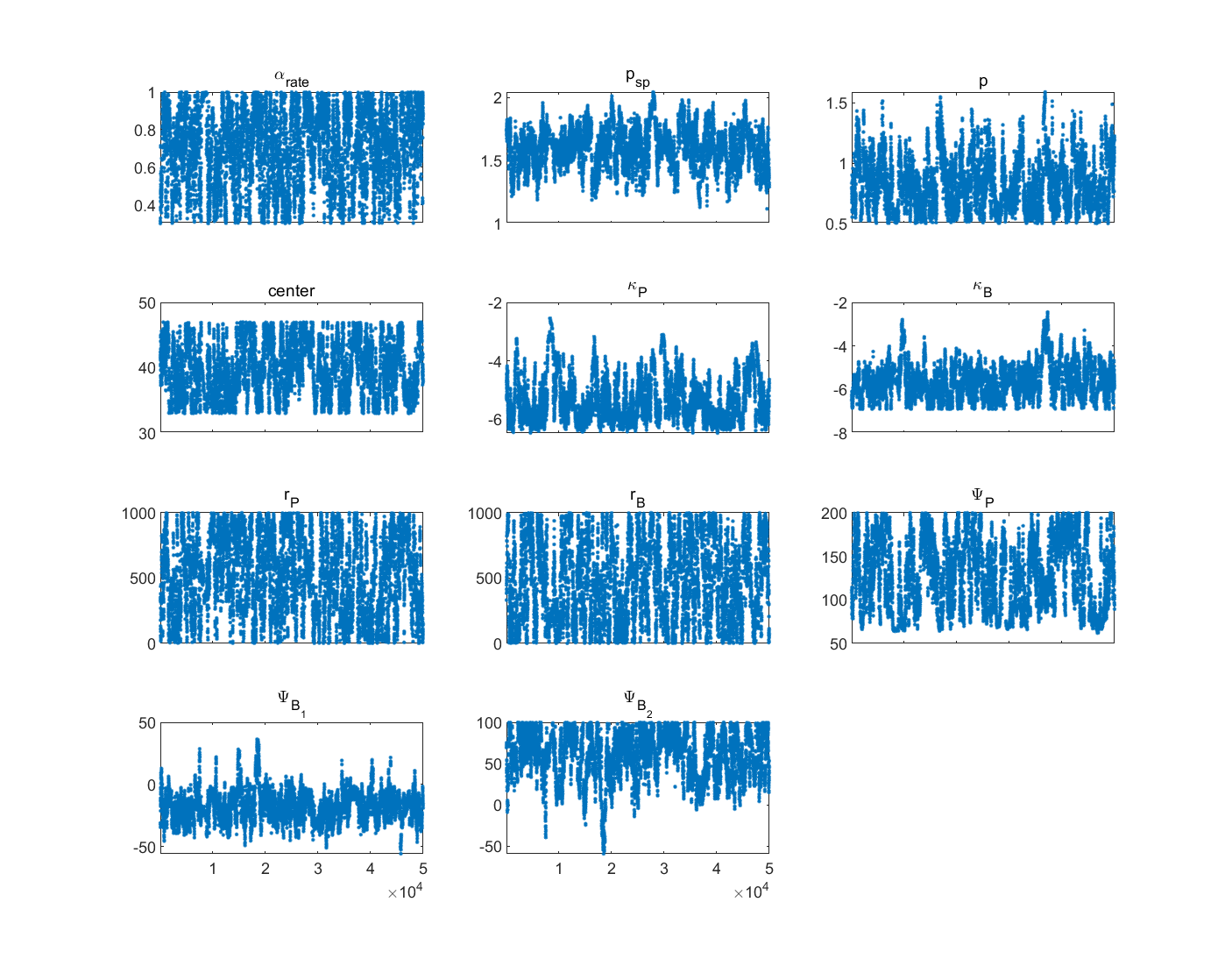

Supplementary Figure 10. Chain-panel plot during the first segment (Nov. 11, 2021 – Feb. 26, 2022) in Phase 3.

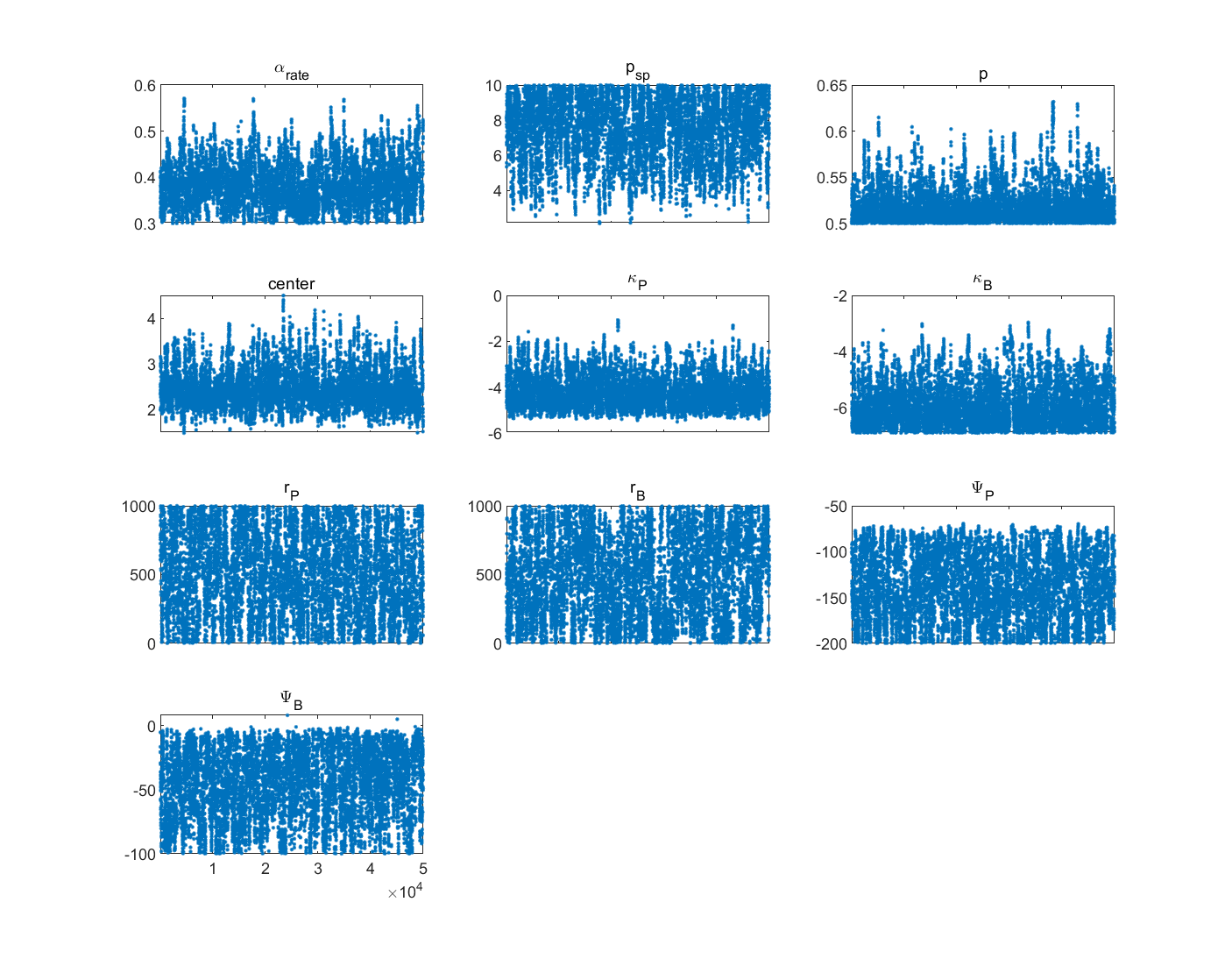

Supplementary Figure 11. Chain-panel plot during the second segment (Feb. 26, 2022 – May. 23, 2022) in Phase 3.

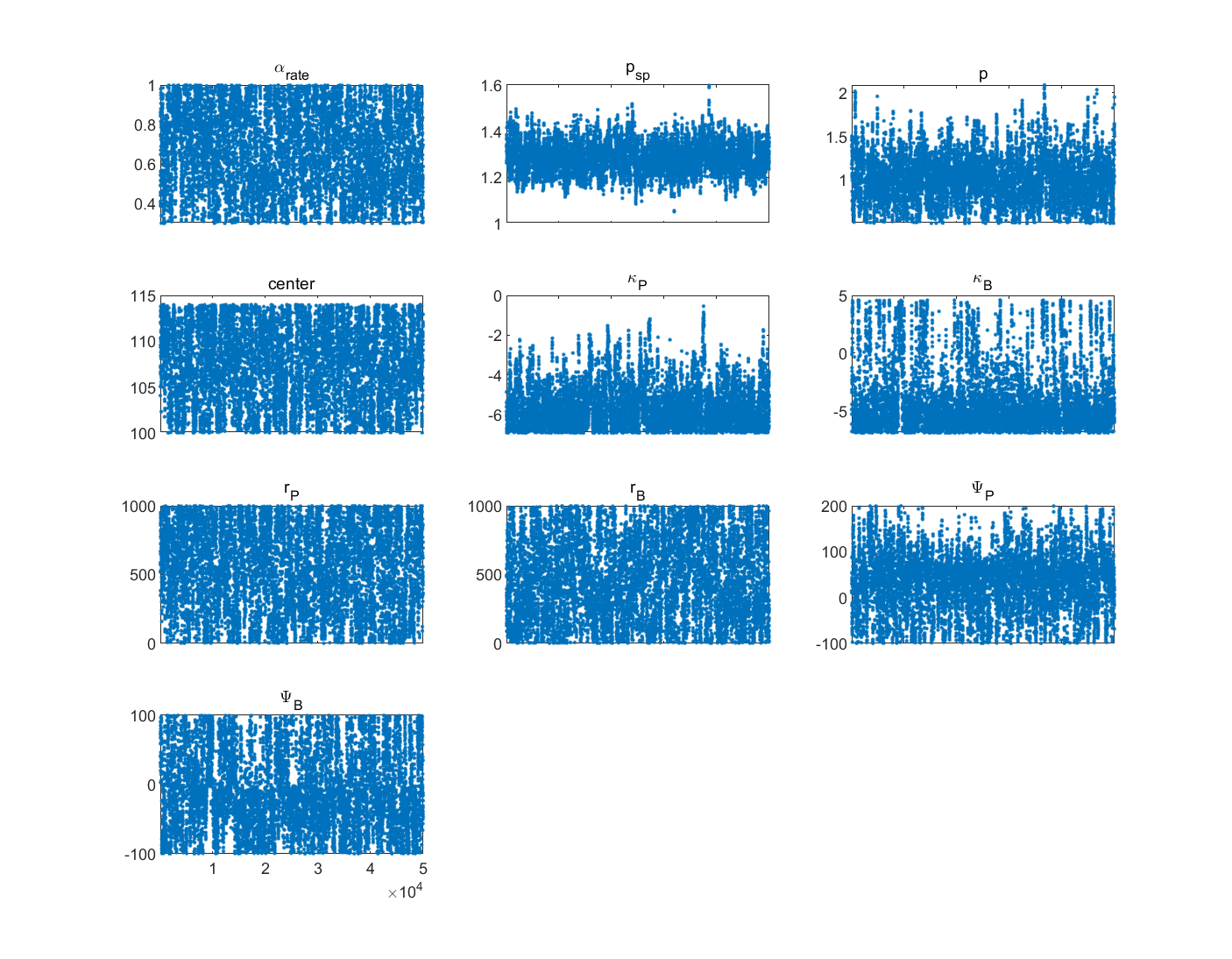

Supplementary Figure 12. Chain-panel plot during the third segment (May. 23, 2022 – Oct. 15, 2022) in Phase 3.

Supplementary Table 3: Fitted parameters using MCMC

| Fitted parameters | Meaning | Phase1-1 | Phase1-2 | Phase1-3 | Phase1-4 | Phase1-5 | Phase2-1 | Phase2-2 | Phase2-3 | Phase3-1 | Phase3-2 | Phase3-3 |
| --- | --- | --- | --- | --- | --- | --- | --- | --- | --- | --- | --- | --- |
| $p_{sp}$ | The adjustment coefficient for superspreading | 5.56  (4.76,6.36) | 2.70  (2.30,3.10) | 8.63  (7.83,9.42) | 2.73  (2.55,2.92) | 1.64  (1.44,1.84) |  |  |  | 1.63  (1.44,1.82) | 7.21  (5.44,8.98) | 1.29  (1.22,1.35) |
| $p$ | The adjustment coefficient for implementing PHSMs or low infection | 1.14  (1.00,1.29) | 0.77  (0.64,0.89) | 1.00  (0.92,1.07) | 1.20  (1.13,1.27) | 0.87  (0.76,0.98) | 1.10  (0.93,1.28) | 0.64  (0.52,0.76) | 0.65  (0.60,0.69) | 0.84  (0.63,1.06) | 0.52  (0.50,0.55) | 1.02  (0.76,1.28) |
| $center$ | Tipping point of PHSMs’ impact | 14.19  (12.76,15.64) | 23.10  (19.78,26.42) | 22.54  (20.67,24.41) | 31.93  (29.63,34.22) | 19.25  (13.66,24.85) |  |  |  | 40.92  (36.98,44.85) | 2.56  (2.10,3.01) | 107.06  (102.98,111.15) |
| ${center}_{vac}$ | Tipping point of incentive measures’ impact |  |  |  |  |  |  |  | 125.60  (118.66,132.53) |  |  |  |
| $\alpha_{rate}$ | The underreported rate | 0.62  (0.43,0.81) | 0.57  (0.39,0.76) | 0.75  (0.57,0.93) | 0.46  (0.31,0.61) | 0.37  (0.31,0.42) | 0.69  (0.51,0.86) | 0.34  (0.30,0.37) | 0.48  (0.35,0.60) | 0.64  (0.44,0.84) | 0.38  (0.33,0.42) | 0.65  (0.46,0.85) |
| $\kappa^{P}$ | Imitation rate of primary doses vaccination |  |  |  |  |  | 0.012  (0.005,0.026) | 0.013  (0.004,0.040) | 0.006  (0.004,0.008) | 0.005  (0.002,0.011) | 0.017  (0.007,0.042) | 0.005  (0.001,0.019) |
| $\kappa^{B}$ | Imitation rate of booster doses vaccination |  |  |  |  |  |  |  |  | 0.003  (0.002,0.006) | 0.003  (0.001,0.005) | 0.022  (0.001,0.421) |
| $r^{P}$ | The adjustment coefficient for the difference in case fatality rate after primary doses vaccination |  |  |  |  |  | 531  (240,822) | 511  (218,804) | 585  (308,861) | 580  (314,845) | 476  (196,757) | 499  (220.02,778.69) |
| $r^{B}$ | The adjustment coefficient for the difference in case fatality rate after booster dose vaccination |  |  |  |  |  |  |  |  | 565  (289,841) | 498  (210,785) | 483.09  (196.00,770.18) |
| $\Psi_{baseline}^{P}$ | The baseline payoff gain for primary doses under different contexts |  |  |  |  |  | 48.91  (19.08,78.73) |  | 61.66  (43.19,80.12) | 63.86  (21.04,106.68) | -70.47  (-107.04,-33.90) | -37.40  (-100.12,25.33) |
| $\Psi_{baseline}^{B}$ | The baseline payoff gain for booster doses under different contexts |  |  |  |  |  |  |  |  | -19.74  (-29.81,-9.67) | -46.22  (-71.79,-20.65) | -10.71  (-64.93,43.52) |
| $\Psi_{E0}$ | The payoff gain following vaccine packaging defects (E0) |  |  |  |  |  |  | -131.80  (-172.09,-89.51) |  |  |  |  |
| $\Psi_{T1}$ | The payoff gain following serious adverse events after vaccination (T1) |  |  |  |  |  |  |  | -71.92  (-89.17,-54.66) |  |  |  |
| $\Psi_{T2}$ | The payoff gain following government and business incentive measures (T2) |  |  |  |  |  |  |  | 64.63  (38.86,90.40) |  |  |  |
| $\Psi_{T3}$ | The payoff gain following reduced information about deaths possibly linked to vaccination |  |  |  |  |  |  |  | -55.01  (-81.50,-28.52) |  |  |  |
| $\Psi_{T4}$ | The payoff gain following end of the incentive measures |  |  |  |  |  |  |  | -67.52  (-87.87,-47.17) |  |  |  |
| $\Psi_{1}^{B}$ | The payoff gain following vaccination priority groups to be expanded to cover people aged 18 or above |  |  |  |  |  |  |  |  | 65.14  (42.30,87.98) |  |  |

Reference

1.	The Government of the Hong Kong Special Administrative Region. COVID-19 Vaccination Programme. https://www.chp.gov.hk/en/features/106934.html.

2.	The Government of the Hong Kong Special Administrative Region. Third dose COVID-19 vaccination arrangements for persons under certain groups. https://www.info.gov.hk/gia/general/202111/03/P2021110300536.htm?fontSize=1.

3.	Bauch C. T. Imitation dynamics predict vaccinating behaviour. *Proc. R. Soc. B Biol. Sci.* **272**, 1669–1675 (2005).

4.	Jentsch, P. C., Anand, M. & Bauch, C. T. Prioritising COVID-19 vaccination in changing social and epidemiological landscapes: a mathematical modelling study. *Lancet Infect. Dis.* **21**, 1097–1106 (2021).

5.	Census and Statistics Department of the government of Hong Kong Special Administrative Region. Population census 2021. https://www.census2021.gov.hk/tc/census_results.html.

6.	The Government of the Hong Kong Special Administrative Region. Transcript of remarks of press conference on anti-epidemic measures. https://www.info.gov.hk/gia/general/202203/11/P2022031100477.htm?fontSize=1.

7.	Campbell, F. *et al.* Increased transmissibility and global spread of SARS-CoV-2 variants of concern as at June 2021. *Eurosurveillance* **26**, (2021).

8.	Bálint, G., Vörös-Horváth, B. & Széchenyi, A. Omicron: increased transmissibility and decreased pathogenicity. *Signal Transduct. Target. Ther.* **7**, 151 (2022).

9.	Miller, I. F., Becker, A. D., Grenfell, B. T. & Metcalf, C. J. E. Disease and healthcare burden of COVID-19 in the United States. *Nat. Med.* **26**, 1212–1217 (2020).

10.	Nasreen, S. *et al.* Effectiveness of COVID-19 vaccines against symptomatic SARS-CoV-2 infection and severe outcomes with variants of concern in Ontario. *Nat. Microbiol.* **7**, 379–385 (2022).

11.	LKS Faculty of Medicine of The University of Hong Kong. Hkumed updates on modelling the fifth wave of covid- 19 in hong kong. https://www.med.hku.hk/en/news/press/-/media/DF5A2F6918764DC4B6517CE7B5F2796B.ashx.

12.	Buchan, S. A. *et al.* Estimated Effectiveness of COVID-19 Vaccines Against Omicron or Delta Symptomatic Infection and Severe Outcomes. *JAMA Netw. Open* **5**, e2232760 (2022).

13.	Hao, X. *et al.* Reconstruction of the full transmission dynamics of COVID-19 in Wuhan. *Nature* **584**, 420–424 (2020).

14.	Prem, K. *et al.* Projecting contact matrices in 177 geographical regions: An update and comparison with empirical data for the COVID-19 era. *PLOS Comput. Biol.* **17**, e1009098 (2021).
